# The Paradox of Neglecting Changes in Behavior: How Standard Epidemic Models Misestimate Both Transmissibility and Final Epidemic Size

**DOI:** 10.64898/2025.12.07.25341782

**Authors:** Binod Pant, Marko Lalovic, István Z. Kiss, Mauricio Santillana

## Abstract

During epidemic outbreaks, populations adapt their behavior in response to disease burden, fundamentally altering transmission dynamics. Despite this, most compartmental models assume constant contact rates throughout outbreaks. To quantify biases from this assumption, we fitted a baseline SEIRD model with constant transmission and three behavioral variants—incorporating mortality-driven transmission reduction via exponential, rational, and mixed functional forms—to COVID-19 mortality data from 30 US locations during the first pandemic wave (March–July 2020). All three behavioral models achieved a lower median normalized sum of squared error in at least 28 of 30 locations, and Bayesian model selection favored them in at least 28 of 30 locations. More importantly, we identified systematic biases when behavioral responses are ignored: the baseline model consistently underestimated the basic reproduction number (ℛ_0_) while paradoxically overestimating the final epidemic size. Median ℛ_0_ estimates from the behavioral models exceeded the baseline estimates across all 30 locations, yet baseline models predicted larger cumulative infection burdens. Controlled synthetic experiments—where mortality trajectories were generated from behavioral models with known parameters—confirmed these biases result from model misspecification rather than data quality or stochastic variation. We prove analytically that for any fixed ℛ_0_, the baseline model overestimates cumulative infections compared to behavioral models where mortality reduces transmission, regardless of functional form. This dual bias poses serious risks for pandemic response: standard models may simultaneously underestimate pathogen contagiousness (delaying critical early action) while overestimating infection burden (causing excessive late-phase resource allocation). Our findings across 30 geographically diverse locations demonstrate that incorporating behavioral change substantially improves both model fit and estimation of epidemiological parameters essential for public health policy.

**Author Summary:** When diseases spread, people change their behavior—avoiding crowds, wearing masks, washing hands more frequently. Yet most mathematical models used to predict epidemics assume people behave the same way throughout an outbreak. We asked: What happens when models ignore these behavioral changes? Using COVID-19 data from 30 US locations, we compared a traditional model (assuming constant behavior) against models where people reduce contact as deaths increase. We discovered a troubling paradox: models ignoring behavior consistently suggest diseases are less contagious than they really are, yet simultaneously predict that more people will get infected. This creates a dangerous mismatch for decision-makers: underestimating how easily a disease spreads may delay urgent early actions like school closures or travel restrictions, while overestimating total infections wastes resources preparing for scenarios that behavioral adaptation prevents. We confirmed this paradox through computer simulations where we knew the true answer, and proved analytically that behavioral adaptation reduces final epidemic size even when the basic reproduction number is held constant. Our work shows that incorporating human behavioral responses is not just a modeling refinement—it is essential for accurate epidemic predictions that inform life-or-death policy decisions.

## 1 Introduction

Behavioral responses to epidemic threats fundamentally shape disease transmission. During the 1918 influenza pandemic, communities closed schools, banned public gatherings, and adopted mask-wearing to reduce transmission [1, 2]. The HIV/AIDS epidemic saw widespread adoption of condom use, averting an estimated 100 million infections between 1990 and 2019 [3]. More recently, the COVID-19 pandemic triggered rapid behavioral shifts, including masking and social distancing, with mask mandates alone associated with significant reductions in cases and deaths [4, 5]. Recent survey data from the United States suggests that human behavior, which varied spatially and temporally during COVID-19, was strongly associated with changes in COVID-19 mortality and hospitalization [6], indicating that observed disease severity indicators drive behavioral adaptation. These examples demonstrate that human populations do not passively experience epidemics—they adapt their behavior in response to perceived risk, fundamentally altering transmission dynamics.

Compartmental models based on Susceptible-Exposed-Infected-Recovered (SEIR) frameworks have been widely used to understand epidemic trajectories and guide public health interventions [7]. Despite historical and contemporary evidence showing that people actively adapt their behavior in response to disease threats, most models assume populations passively experience outbreaks by maintaining constant contact patterns. Although the assumption of constant transmission rates is mathematically convenient, it ignores the dynamic feedback between disease burden and behavioral change. Recent modeling efforts have begun incorporating behavioral responses through various mechanisms, including prevalence-dependent [8–11], hospitalization-driven [12], and mortality-driven feedback [8, 13, 14]. Lejeune et al. [15] classify these approaches as either endogenous (behavior changes as a function of disease state variables within the model, creating a feedback loop) or exogenous (behavior changes through fixed or time-varying parameters informed by external data sources such as government policies and mobility patterns, or through modelers’ assumptions, rather than a state variable within a model itself).

The critical question is whether systematic biases arise when models fail to account for behavioral change. If populations adapt their behavior in response to disease burden—as historical and contemporary evidence suggests—then models assuming constant transmission may systematically mischaracterize epidemic dynamics. Such mischaracterization could have serious consequences for public health decision-making: underestimating pathogen transmissibility might lead to insufficient interventions, while overestimating infection burden could result in misallocated resources. Despite these stakes, few studies have systematically investigated—either numerically or analytically—whether and to what extent omitting behavioral feedback introduces biases in epidemic models. Recent work has demonstrated that incorporating behavioral responses improves model fit to observed data [8, 12, 14, 16, 17], with behavioral models capturing epidemic trajectories better than their behavior-free counterparts. Behavioral models have also shown superior performance in out-of-sample validation [8, 12]. Theoretical studies have examined how behavioral mechanisms—including vaccination behavior [18], awareness-driven protection [19], and incidence-dependent contact reduction [9]— alter epidemic outcomes such as cumulative infection burden; notably, models ignoring behavior change have systematically overshoot final size projections [20]. However, systematic comparison of key epidemiological quantities—such as basic reproduction number and final size—estimated through models with versus without behavioral feedback, using real data across many locations, remains lacking.

We address this gap by systematically comparing compartmental models with and without behavioral feedback, fitted to COVID-19 mortality data from 30 US locations (29 states and the District of Columbia) during the first pandemic wave (March–July 2020). We implement three behavioral models where the transmission rate decreases as mortality increases, alongside a baseline model with constant transmission. Using Bayesian methods, we fit each model to observed mortality trajectories, estimate posterior distributions for the basic reproduction number and final epidemic size, and compare model performance both qualitatively and quantitatively. To test whether biases arise from model misspecification rather than data limitations, we conduct controlled synthetic experiments where true parameter values are known by design. We also analytically derive the relationship between final epidemic size in baseline versus behavioral models when the basic reproduction number is identical.

Our analysis reveals three key findings with important public health implications. First, behavioral models consistently outperform baseline models across nearly all locations examined, as evidenced by both qualitative and quantitative comparisons. Second, baseline models systematically underestimate the basic reproduction number while counterintuitively overestimating the total proportion of the population infected compared to all three behavioral model formulations. We confirm this dual bias through synthetic experiments, demonstrating that these biases emerge when behavioral feedback is present in the data but models do not explicitly account for behavioral change. Third, we prove analytically that this paradox—lower estimated transmissibility yet higher predicted final epidemic size—is an inevitable consequence of ignoring behavioral responses. These findings demonstrate that incorporating behavioral dynamics is not merely a modeling refinement but essential for accurate epidemic parameter estimation and evidence-based pandemic response.

This paper is organized as follows. In Section 2, we formulate the baseline model that does not explicitly account for behavioral change and three behavioral model variants, describe the COVID-19 mortality data used for calibration, and outline our Bayesian parameter inference approach. Section 3 presents our three key findings: behavioral models’ superior fit to observed data, systematic underestimation of the basic reproduction number by baseline models, and overestimation of infection burden when behavioral responses are ignored. We support these findings through both real data analysis across 30 US states and controlled synthetic experiments. Section 4 discusses the public health implications and limitations of our work.

## 2 Methods

### 2.1 Model Formulation

We developed a compartmental SEIRD model incorporating behavioral responses to disease-induced mortality. The population is stratified into susceptible (*S*), exposed (*E*), infectious (*I*), recovered (*R*), and deceased (*D*) compartments, with total living population *N* (*t*) = *S*(*t*) + *E*(*t*) + *I*(*t*) + *R*(*t*) at time *t*. The model dynamics are governed by the following system of differential equations:

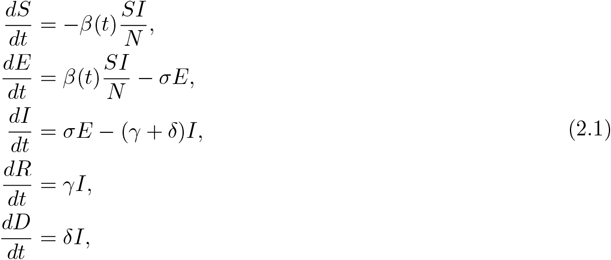

where *σ* is the incubation rate, *γ* is the recovery rate, and *δ* is the disease-induced mortality rate. The transmission rate *β*(*t*) varies between model variants to capture different behavioral assumptions, whereby the severity of the outbreak is assumed to lead to behavioral adaptation aimed at reducing infection risk.

#### Baseline model

The baseline model assumes constant transmission *β*(*t*) = *β*_0_, representing traditional SEIRD dynamics without explicit behavioral feedback.

#### Behavioral models

Behavioral models incorporate dynamic feedback through mortality-dependent transmission, with *β*(*t*) declining as deaths increase to reflect behavioral adaptation to perceived risk. The transmission rate *β*(*t*) depends on three quantities: *β*_0_, the baseline transmission rate in the absence of behavioral response; *δI*(*t*), the instantaneous death rate due to disease; and *ζ*, which quantifies the population’s sensitivity to mortality. Larger values of *ζ* correspond to stronger behavioral responses. We consider three functional forms of *β*(*t*):

a. *Exponential form:*

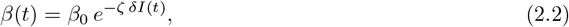

where transmission rate decays exponentially with increasing mortality.
b. *Rational form:*

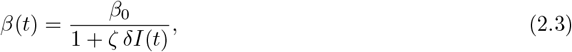

which produces similar behavior to the exponential form but saturates more slowly as deaths rise.
c. *Mixed form:*

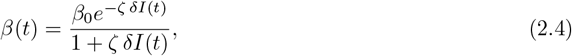

which incorporates both exponential and rational forms. This hybrid form saturates faster than either individual form as mortality increases.

All behavioral models reduce to the baseline model when *ζ* = 0. Hereafter, we refer to these as the *Behavior (Exponential), Behavior (Rational)*, and *Behavior (Mixed)* models.

#### Daily mortality

While individuals change behavior in response to instantaneous mortality *δI*, we compare model predictions to observed daily mortality data. Model-simulated daily mortality on day *t* is defined as:

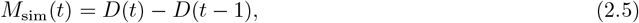

representing the daily increment in cumulative deaths. The time series *M*_sim_(*t*)_*t*_ is compared to observed mortality data to estimate model parameters (Section 2.3).

#### Basic reproduction number

The *basic reproduction number*, defined as the average number of secondary infections produced by a typical infectious individual in a wholly susceptible population (i.e., *S*(0) ≈ *N* (0)), is given by:

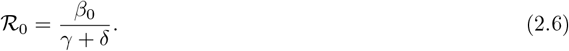

#### Effective reproduction number

The *effective reproduction number*, denoted by ℛ_*e*_(*t*), is the average number of new infections generated by a typical infected individual at time *t* [21]. The effective reproduction numbers for the baseline model, 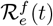, and behavioral model, 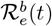, are given respectively by:

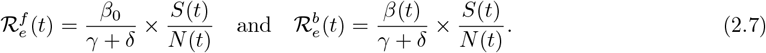

#### Final epidemic size

The final epidemic size, denoted *A*, represents the total proportion of the population that becomes infected over the course of an outbreak:

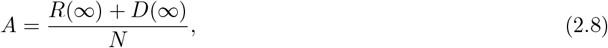

where *R*(∞) and *D*(∞) denote the cumulative recovered and deceased individuals as *t* → ∞. Numerically, we compute *A* from simulated SEIRD trajectories by integrating until the infectious compartment falls below a threshold (*I*(*t*) *<* 1) and calculating *A* = (*N* − *S*(*t*_end_))*/N*, where *t*_end_ is the time at which the epidemic effectively terminates.

### 2.2 Data and State Selection

We calibrated the models to COVID-19 mortality data from the COVID-19 Data Repository by the Center for Systems Science and Engineering (CSSE) at Johns Hopkins University [22, 23]. Our analysis focused on the first wave of the pandemic in the United States, covering the 123-day period from March 1, 2020 to July 1, 2020.

#### State selection criteria

From the 51 states and territories in the dataset, we selected 30 locations for analysis based on three criteria designed to ensure data quality and the presence of a well-defined epidemic wave:

1. *Sufficient mortality signal:* States with very low daily death counts produced noisy 7-day moving averages unsuitable for reliable parameter inference (excluded: AK, HI, ID, ME, MT, ND, OR, SD, UT, VT, WV, WY).
2. *Clear single-wave structure:* States must exhibit a distinct epidemic wave with a recognizable peak and subsequent decline within the observation window (excluded: AL, AR, AZ, CA, FL, SC, TN, TX).
3. *Data integrity:* States with apparent reporting anomalies, such as sudden discontinuities in cumulative deaths, were excluded (excluded: CO).

The 30 selected locations were: Connecticut, Delaware, District of Columbia, Georgia, Illinois, Indiana, Iowa, Kansas, Kentucky, Louisiana, Maryland, Massachusetts, Michigan, Minnesota, Mississippi, Missouri, Nebraska, New Hampshire, New Jersey, New Mexico, Nevada, New York, North Carolina, Ohio, Oklahoma, Pennsylvania, Rhode Island, Virginia, Washington, and Wisconsin. Details on the 21 excluded locations and their mortality trajectories are provided in Supplementary Figure S1.

#### Data preprocessing

We applied a trailing 7-day moving average to daily mortality counts to reduce reporting artifacts.

#### Population data

Initial state population sizes *N* (0) were obtained from the JHU CSSE dataset, ranging from 705,749 (District of Columbia) to 19,453,561 (New York).

### 2.3 Parameter Inference

We infer parameters using Approximate Bayesian Computation with Sequential Monte Carlo (ABC–SMC) [24], as implemented in the pyABC library [25], with the deterministic SEIRD model (2.1). The complete codebase for simulation, inference, and figure generation is available at https://github.com/markolalovic/epibehavior-models [26]. For each location and model, we simulate daily deaths over the 123-day window and apply the same 7-day trailing average used for the observed data.

We reparameterize in epidemiological terms, inferring ℛ_0_, the initial prevalence *π*_0_ = *I*_0_*/N*, mortality rate *δ*, and behavioral response parameter *ζ*, with weakly informative priors:

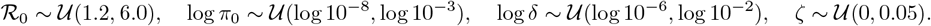

ABC–SMC uses 1000 particles per generation, a multivariate normal transition kernel adapted from the previous population, and a quantile-based tolerance schedule over up to 10 generations. Additional details are provided in Supplement Section S2.

### 2.4 Synthetic Data Experiments

To rigorously test whether behavioral effects can be recovered from mortality data and to quantify systematic biases in parameter estimation, we conducted controlled synthetic data experiments. We generated mortality trajectories from behavioral models using parameters calibrated to Massachusetts COVID-19 data, with varying values of the behavioral parameter *ζ*. To approximate observational variability, we added small independent Gaussian noise to each simulated series:

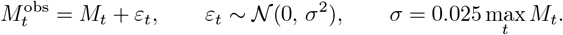

Negative values were truncated to zero before applying the 7-day moving average. We then fitted both the data-generating behavioral model and the baseline model to these synthetic mortality datasets. This approach allows us to assess: (i) whether the behavioral model correctly recovers the known underlying dynamics; (ii) whether the baseline model produces systematic biases when behavioral feedback is present but ignored; and (iii) how the magnitude of behavioral response (controlled by varying *ζ*) affects the ability to distinguish between model structures. By knowing the true data-generating mechanism, we can definitively attribute any discrepancies in estimation of ℛ_0_ and final epidemic size to model misspecification rather than data limitations or stochastic variation. Additional details are provided in Supplement Section S3.

### 2.5 Model Comparison

We evaluate the relative performance of baseline and behavioral models using Bayesian model selection via ABC–SMC. We describe below the distance metric and Bayes factor calculation used for model comparison.

#### Distance metric

To quantify the discrepancy between model-simulated and observed mortality trajectories, we use the *Normalized Sum of Squared Errors* (NSSE):

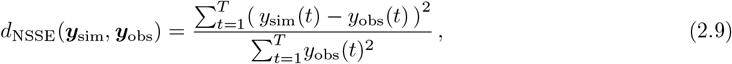

where *y*_sim_(*t*) is the model-simulated daily mortality time series and *y*_obs_(*t*) is the observed 7-day averaged daily mortality series over the *T* = 123 day observation window.

The NSSE is the sum of squared errors normalized by a constant that depends only on the observed data for each location. This normalization makes the distance scale-invariant across locations with different mortality burdens, enabling comparable tolerance thresholds and performance metrics across all states while preserving the same ordering of parameter sets that would arise from the unnormalized sum of squared errors within any given state.

#### Bayes factor

We performed ABC–SMC model selection [24] comparing the baseline model against each of the three behavioral model formulations (exponential, rational, and mixed) separately, assigning equal prior probabilities to each pair. To avoid extinction artifacts, we report results at the first stabilized generation *g*^∗^, defined as the earliest *g* for which both model probabilities change by less than 0.02 over 3 consecutive generations and *P* (Baseline |*D, g*) *>* 0. At the stabilized generation *g*^∗^, we summarize the evidence for a behavioral model via the Bayes factor under equal model priors:

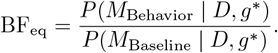

We interpret BF_eq_ following the scale from [27]. Values of BF_eq_ *>* 1 indicate evidence favoring the behavioral model over the baseline model, with strength categorized as: 1–3 (*very weak*), 3–20 (*positive*), 20–150 (*strong*), and *>* 150 (*very strong*). Values *<* 1 indicate evidence favoring the baseline model.

## 3 Results

We fitted COVID-19 mortality data from 30 US locations (29 states and the District of Columbia) during the first pandemic wave (March–July 2020) to a baseline SEIRD model with constant transmission rate and three behavioral SEIRD models where transmission rate declines in response to disease-induced mortality. We selected these 30 locations from the 50 states and DC based on data quality criteria including sufficient mortality signal, clear single-wave structure, and data integrity (see Section 2 for detailed selection criteria). Our analysis reveals three key findings: (i) behavioral models provide significantly improved fits to observed mortality data across almost all locations examined, with both median normalized squared error and Bayes factors favoring behavioral models in 28 of 30 locations; (ii) baseline models that ignore behavioral change systematically underestimate the basic reproduction number (*R*_0_), with median *R*_0_ from behavioral models consistently higher than from baseline models across all states; and (iii) baseline models overestimate total infection burden despite underestimating transmissibility, creating a paradoxical dual bias with serious policy implications.

### 3.1 Behavioral Models Show Superior Fit to COVID-19 Mortality Data

All three behavioral models (exponential, rational, and mixed) consistently outperformed the baseline model when fitted to COVID-19 mortality data from 30 locations during the first pandemic wave. Visually, the behavioral models capture the characteristic asymmetric mortality curves observed in many locations—sharp initial rises followed by gradual, extended declines—while the baseline model produces curves that decay too rapidly (Figure 1 shows the mixed behavioral model; Supplementary Figures S4 and S5 show exponential and rational formulations, respectively).

**Figure 1.**
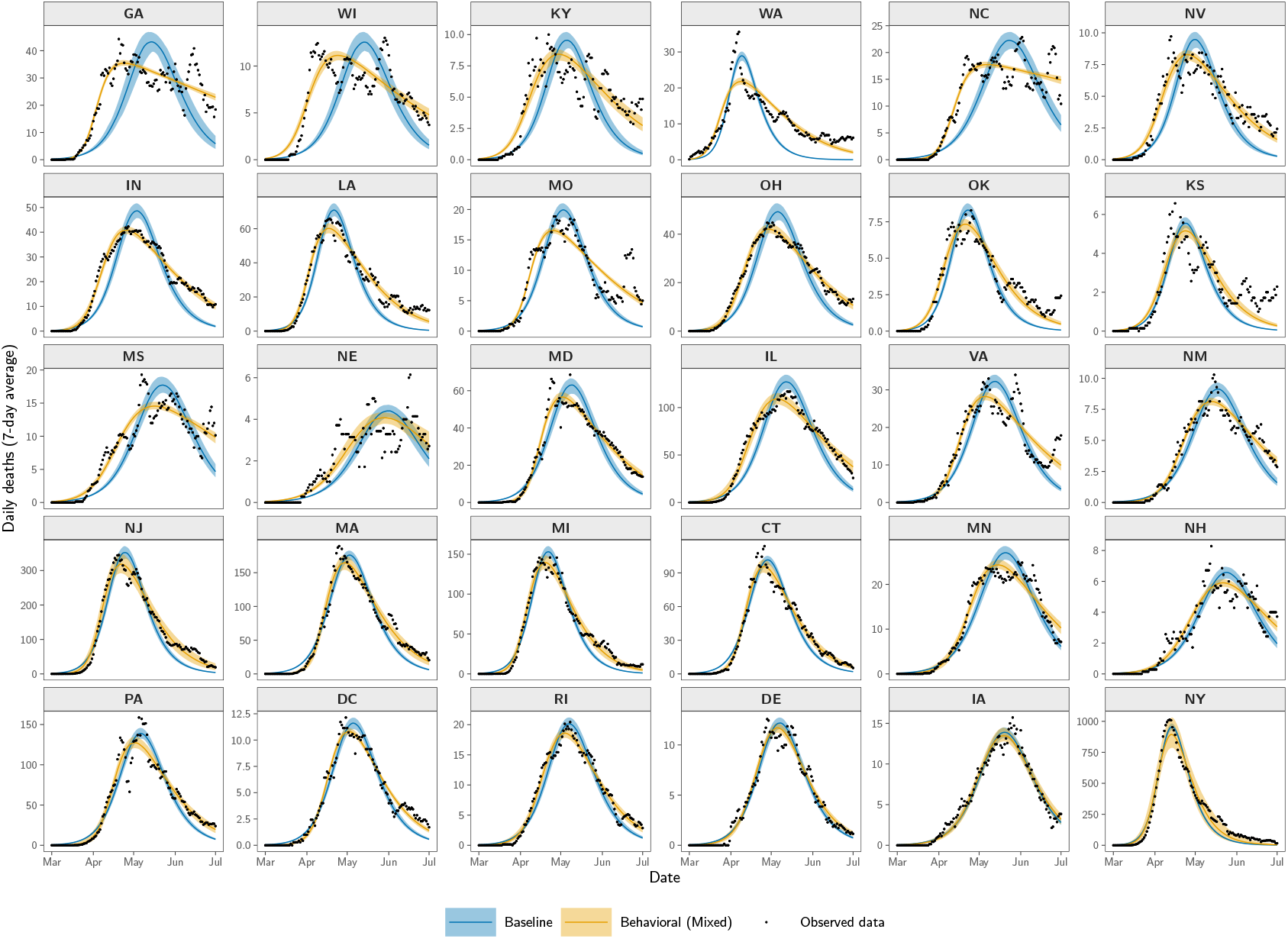
Behavioral models capture observed mortality dynamics better than baseline models across 30 US locations. Posterior predictive fits comparing baseline and behavioral (mixed) models to 7-day averaged daily COVID-19 deaths during the first pandemic wave (March–July 2020). Black points show observed mortality data. Solid lines show posterior predictive medians and shaded regions show 90% credible intervals (5th–95th percentiles), computed from 1,000 weighted posterior samples drawn from the final ABC–SMC generation. The behavioral model accurately captures the characteristic asymmetric mortality curves with sharp initial rises and gradual extended declines observed across most locations. States are ordered from left to right and top to bottom by decreasing improvement in fit: those where the behavioral model outperforms the baseline model most dramatically (largest difference in median NSSE) appear first, progressing to states where the two models perform most similarly. This ordering visually emphasizes that behavioral feedback provides substantial improvements in model fit across the vast majority of locations examined.

This superior performance is confirmed quantitatively across all three formulations. For the mixed behavioral model, we observed lower median normalized sum of squared errors (NSSE) in 28 of 30 locations, with Bayesian model selection via ABC–SMC providing evidence for the mixed behavioral model in these same 28 locations (Table 1). Only Iowa and New York favored the baseline model. The exponential and rational formulations of behavioral models demonstrated comparable performance (see Supplementary Tables S1 and S2). Overall, across all three behavioral formulations, at least 28 of 30 locations favored behavioral models over baseline models based on NSSE and Bayes factor.

**Table 1.** Comparison of the baseline model and the behavioral (mixed) model across 30 U.S. states. For each state, Median NSSE_B_ and Median NSSE_M_ denote the median posterior normalized sum of squared errors (NSSE) under the baseline *B* and behavioral (mixed) *M* models, respectively. BF_eq_ is the Bayes factor under equal model priors in favor of the mixed model versus the baseline model at the stabilized ABC–SMC generation, and “Evidence” gives the corresponding verbal strength of evidence. See Section 2 for interpretation guidelines for BF_eq_.

| State | Median NSSE<sub>B</sub> | Median NSSE<sub>M</sub> | BF<sub>eq</sub> | Evidence |
| --- | --- | --- | --- | --- |
| DC | 0.0285 | **0.0131** | 11551.24 | very strong |
| MS | 0.0882 | **0.0337** | 5051.63 | very strong |
| NH | 0.0571 | **0.0381** | 2105.67 | very strong |
| NE | 0.1390 | **0.0859** | 1852.99 | very strong |
| RI | 0.0250 | **0.0111** | 852.43 | very strong |
| NM | 0.0488 | **0.0151** | 688.87 | very strong |
| MO | 0.1214 | **0.0539** | 211.95 | very strong |
| KY | 0.2154 | **0.0586** | 210.55 | very strong |
| NC | 0.1440 | **0.0310** | 164.63 | very strong |
| IN | 0.0896 | **0.0086** | 126.43 | strong |
| CT | 0.0330 | **0.0121** | 121.80 | strong |
| WI | 0.2540 | **0.0558** | 105.04 | strong |
| VA | 0.0805 | **0.0387** | 100.58 | strong |
| MN | 0.0392 | **0.0188** | 79.31 | strong |
| GA | 0.2254 | **0.0265** | 66.31 | strong |
| WA | 0.1786 | **0.0655** | 64.52 | strong |
| OH | 0.0690 | **0.0060** | 37.48 | strong |
| IL | 0.0572 | **0.0110** | 35.61 | strong |
| OK | 0.1005 | **0.0394** | 28.71 | strong |
| MA | 0.0374 | **0.0145** | 22.96 | strong |
| PA | 0.0471 | **0.0311** | 19.86 | positive |
| LA | 0.0842 | **0.0165** | 13.46 | positive |
| MD | 0.0621 | **0.0151** | 11.97 | positive |
| MI | 0.0344 | **0.0123** | 11.80 | positive |
| DE | 0.0210 | **0.0167** | 10.67 | positive |
| NJ | 0.0433 | **0.0157** | 3.67 | positive |
| NV | 0.1456 | **0.0359** | 1.47 | very weak |
| KS | 0.1627 | **0.1054** | 1.34 | very weak |
| IA | **0.0141** | 0.0169 | 0.51 | favours Baseline |
| NY | **0.0277** | 0.0316 | 0.02 | favours Baseline |

The superior performance of behavioral models varies systematically across locations, with the largest improvements in states exhibiting extended mortality decline phases. Figure 1 orders states by decreasing difference in median NSSE between baseline and behavioral models, with locations showing the largest improvement appearing first. States like Washington and Indiana (top left) exhibit the most striking contrast— the behavioral models capture gradual, extended mortality declines that baseline models fail to reproduce. Moving toward Rhode Island and Delaware (bottom right), the performance gap narrows, with both model types achieving similar fit quality.

Sensitivity analysis reveals a key feature of behavioral models: increasing the behavioral response parameter (*ζ*) produces lower peak mortality and slower decline in daily deaths following the peak (Supplementary Figure S8), a mechanism absent in baseline models. This pattern is reminiscent of the “flattening the curve” phenomenon, where public health interventions reduce peak mortality while extending the duration of the outbreak. In all three behavioral formulations, *β*(*t*) exhibits a characteristic pattern: sharp decline during the mortality surge followed by gradual recovery as deaths decline (see Supplementary Section S6). This time-varying transmission rate better reflects the reality of behavioral adaptation during epidemics, consequently producing superior fits to observed mortality data compared to the constant transmission rate assumption of the baseline model.

We validated this finding through controlled synthetic data experiments where mortality trajectories were generated from behavioral models with varying levels of behavioral sensitivity (*ζ*), then fitted to both the data-generating behavioral model and the baseline model. Using the mixed behavioral formulation, behavioral models accurately recovered the true dynamics across all *ζ* values, while the baseline model increasingly failed to capture mortality trajectories as behavioral feedback strengthened (Figure 3A). Results were consistent across all three functional forms examined (Supplementary Figures S2A and S3A show exponential and rational forms, respectively), demonstrating that baseline models systematically mischaracterize epidemic trajectories when behavioral responses are present.

### 3.2 Baseline Models Systematically Underestimate the Basic Reproduction Number

Baseline models that ignore behavioral change systematically underestimate the basic reproduction number (ℛ_0_) compared to all three behavioral models when fitted to the same COVID-19 mortality data across 30 US locations (Figure 2A shows comparison with mixed formulation). Furthermore, ℛ_0_ estimates from all three behavioral model variants are similar to each other, and the systematic underestimation by baseline models is consistent across exponential, rational, and mixed formulations (Supplementary Figures S6 and S7 show ℛ_0_ comparisons with exponential and rational forms, respectively).

**Figure 2.**
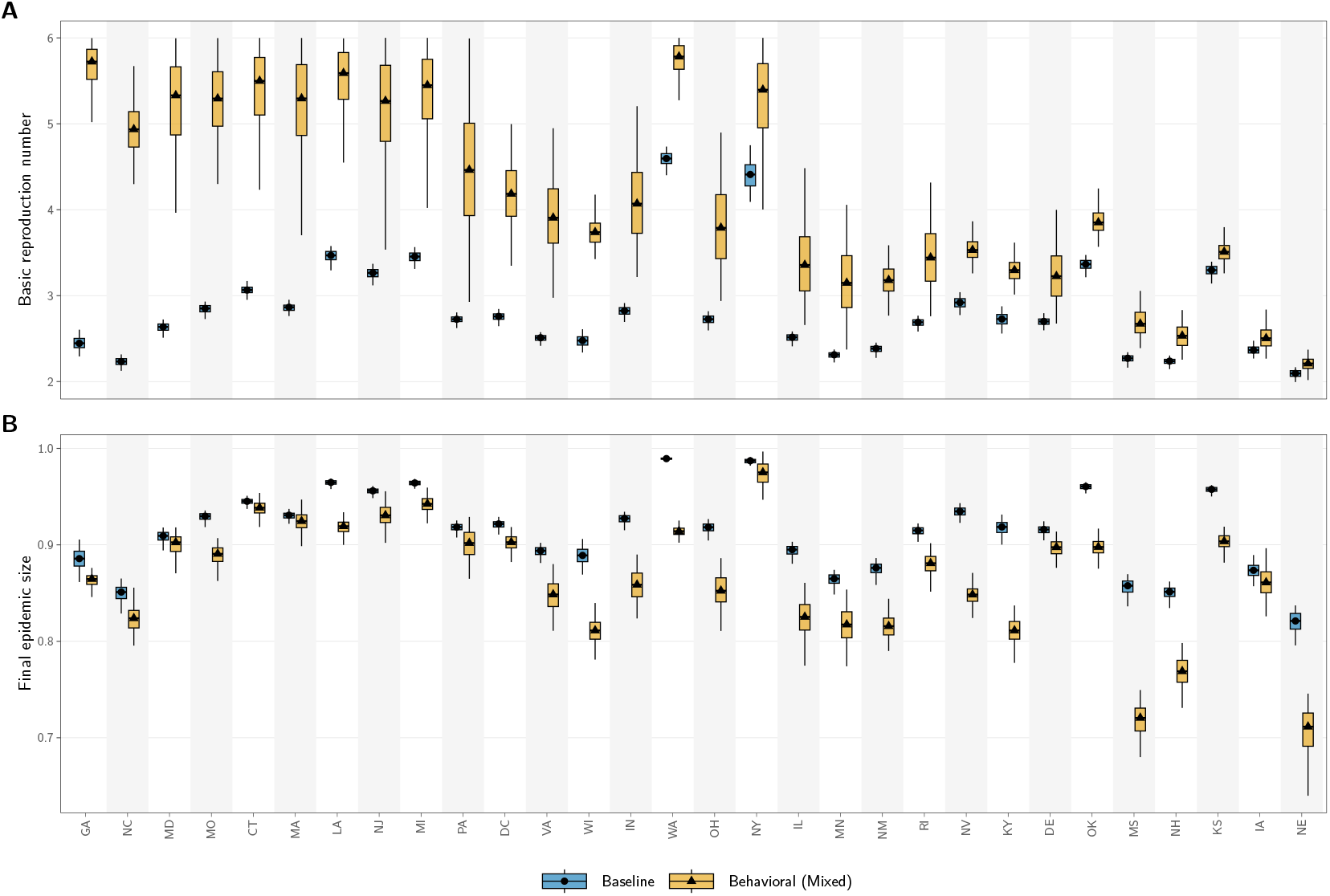
Baseline models systematically underestimate basic reproduction number while overestimating final epidemic size across all 30 locations. Posterior distributions of key epidemiological quantities estimated by fitting the baseline and behavioral (mixed) models to COVID-19 mortality data from 30 US locations during the first pandemic wave (March–July 2020). **(A)** Basic reproduction number ℛ_0_. The behavioral model consistently estimates higher ℛ_0_ values than the baseline model across all 30 locations, indicating that models ignoring behavioral feedback systematically underestimate intrinsic transmissibility. **(B)** Final epidemic size (proportion of population infected). Despite estimating lower ℛ_0_, the baseline model predicts larger cumulative infection burdens than the behavioral model in most locations, demonstrating the paradoxical consequence of ignoring behavioral adaptation. Boxplots show median (center line), interquartile range (box), and whiskers extending to 1.5 *×* IQR within the sample support computed from weighted posterior samples. States are ordered from left to right by decreasing difference in median basic reproduction number between the behavioral (mixed) and baseline models.

The effective reproduction number for all four models is provided in Supplementary Section S7. In all cases, ℛ_*e*_(*t*) converges to values below 1 by late April–May across all states (coinciding with peak infection), though behavioral models exhibit steeper initial declines corresponding to rapid behavioral responses during the early stages of the outbreak.

Synthetic experiments where true ℛ_0_ was known by design confirmed this systematic underestimation. Baseline models consistently underestimated true ℛ_0_ across all behavioral sensitivity values, with bias increasing as behavioral feedback strengthened (increasing *ζ*), while behavioral models accurately recovered true ℛ_0_ across all *ζ* values (Figure 3B for mixed; Supplementary Figures S2B and S3B for exponential and rational forms). This confirms that ℛ_0_ underestimation is a systematic consequence of model misspecification.

**Figure 3.**
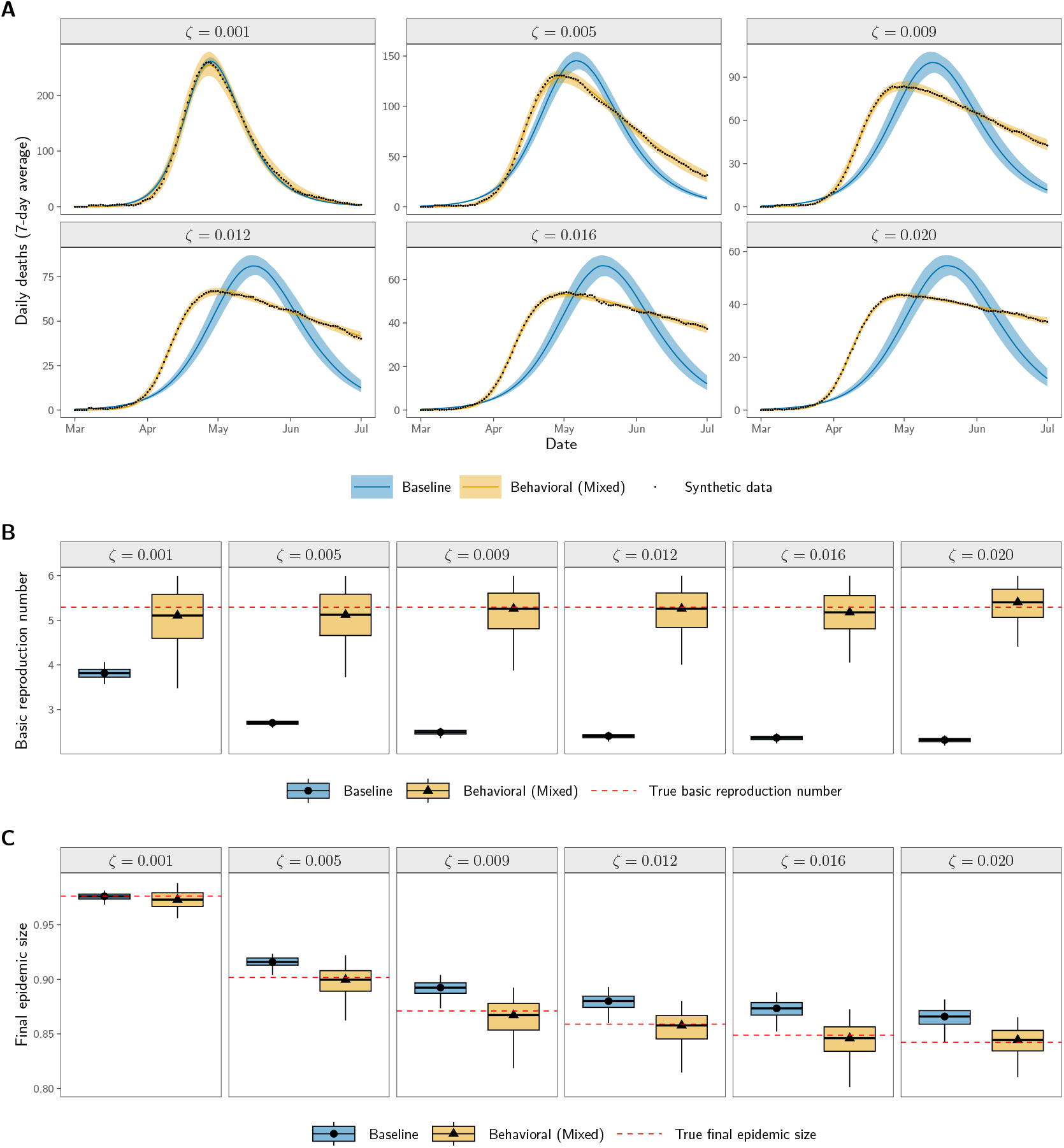
Baseline models systematically misestimate epidemic quantities when behavioral feedback is present but ignored. Synthetic data experiments where mortality trajectories were generated using the behavioral (mixed) model with known parameters and varying behavioral sensitivity *ζ*, then fitted with both baseline and behavioral models. **(A)** Posterior predictive fits to synthetic mortality data (black points). Solid lines show posterior medians and shaded regions show 90% credible intervals. The behavioral (mixed) model accurately recovers the true dynamics across all *ζ* values, while the baseline model increasingly fails to capture the extended post-peak decline as behavioral feedback strengthens with increasing *ζ* values. **(B)** Posterior distributions of the basic reproduction number ℛ_0_. The behavioral model correctly recovers the true ℛ_0_ (red dashed line) across all scenarios, while the baseline model systematically underestimates ℛ_0_, with bias increasing as *ζ* increases. **(C)** Posterior distributions of final epidemic size. The behavioral model accurately estimates the true final size (red dashed line), while the baseline model systematically overestimates infection burden despite underestimating transmissibility.

Intuitively, this systematic underestimation of ℛ_0_ arises because baseline and behavioral models, although fitted to the same observed mortality data, assume fundamentally different transmission mechanisms. The baseline model uses a constant transmission rate *β*_0_ throughout the epidemic, fitting the data with a moderate value that remains fixed. In contrast, behavioral models assume a time-varying transmission rate *β*(*t*) that declines as people adapt their behavior in response to rising mortality. To reproduce the same initial epidemic growth observed in the data, behavioral models require a substantially higher initial transmission rate *β*_0_—which then declines as behavioral responses take effect. As evident from Equation (2.6), ℛ_0_ is an increasing function of *β*_0_; hence models that estimate higher values of *β*_0_ necessarily yield higher estimates of ℛ_0_. Thus, behavioral models, which require higher *β*_0_ to account for subsequent behavioral adaptation, naturally produce higher ℛ_0_ estimates than baseline models. This higher initial transmission rate captured by behavioral models represents the intrinsic transmissibility of the pathogen before any behavioral adaptation occurs. The baseline model, lacking this behavioral feedback mechanism, systematically underestimates this initial transmissibility because it interprets the entire epidemic trajectory—including periods when behavior has already adapted—through the lens of a single, constant transmission rate.

### 3.3 Ignoring Behavioral Responses Overestimates Infection Burden

Despite underestimating ℛ_0_, baseline models overestimate the total proportion of the population that becomes infected (Figure 2B shows comparison with mixed; Supplementary Figures S6B and S7B show comparisons with exponential and rational forms, respectively). This overestimation is most pronounced in locations like Illinois, where baseline models predict median final epidemic sizes over 10 percentage points higher than behavioral models. The three behavioral variants produce similar final size estimates across all 30 locations, while baseline models consistently overestimate infection burden compared to all three behavioral models.

Synthetic experiments confirmed this systematic overestimation of final epidemic size. Baseline models consistently overestimated true final epidemic sizes while behavioral models accurately recovered them across all *ζ* values considered (Figure 3C shows comparison with mixed; Supplementary Figures S2C and S3C show comparisons with exponential and rational forms, respectively). The bias in overestimating final epidemic size increases with stronger behavioral responses, demonstrating that ignoring behavioral dynamics leads to inflated infection burden estimates regardless of functional form.

To gain theoretical insight into the relationship between behavioral feedback and epidemic outcomes, we analytically examine final epidemic size under a controlled scenario where ℛ_0_ is identical for both the baseline and a behavioral model. For analytical tractability, we assume a constant total population *N* = *S* +*E* +*I* +*R*+*D*, which differs from the time-varying living population *N* (*t*) = *N* (0) − *D*(*t*) used in our numerical simulations. We prove that for any fixed value of ℛ_0_, behavioral models incorporating mortality-driven feedback produce final epidemic sizes less than or equal to those from baseline models. We first demonstrate this result for the exponential behavioral model.

#### Theorem 3.1

(Final Size Reduction: Exponential Form). *Consider an epidemic model with behavioral response function β*(*t*) = *β*_0_ exp(−*αI*(*t*)) *where α* = *ζδ. Furthermore, we assume that the total population size is fixed with N* = *S* + *E* + *I* + *R* + *D. Let A*_0_ *denote the final size of the baseline model (i*.*e*., *α* = 0*). The final size of the behavioral model, A*_*α*_, *can be expressed as:*

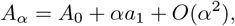

*where a*_1_ *<* 0 *is the first-order correction to the final epidemic size due to behavioral feedback. Hence, for small α, the final size of the behavioral model is smaller than that of the baseline model*.

The proof of Theorem 3.1, based on asymptotic expansion, is given in Appendix A.

This result extends beyond the exponential form. For any behavioral response function satisfying 0 *< β*(*t*) ≤ *β*_0_—including the exponential, rational, and mixed forms considered in this study—we show that behavioral models always produce smaller or equal final epidemic sizes compared to baseline models when ℛ_0_ is the same for both models.

#### Theorem 3.2

(Final Size Reduction: General Form). *Consider the SEIRD model* (2.1) *where N* = *S* + *E* + *I* + *R* + *D is the fixed total population size and the transmission rate β*_*α*_(*t*) = *β*_0_Ψ(*α, I*_*α*_(*t*)) *decreases with increasing mortality through a behavioral response function satisfying* 0 *<* Ψ(*α, I*_*α*_(*t*)) ≤ 1, *with β*_0_ *being the maximum transmission rate and α* = *ζδ. If a behavioral model (with α >* 0*) and the baseline model (with α* = 0 *and constant transmission β*_0_*) have equal basic reproduction numbers, given by* ℛ_0_ = *β*_0_*/*(*γ* + *δ*), *then the final epidemic size of the baseline model (A*_0_*) is always larger than or equal to that of the behavioral model (A*_*α*_).

The proof of Theorem 3.2 is given in Appendix B. We obtain final size relationships for the baseline model with constant transmission rate *β*_0_ and for behavioral models with *β*(*t*) *< β*_0_. By defining an auxiliary function based on these final size equations and exploiting its properties, including its unique maximum and concavity, we show that the final size of the behavioral model is always less than or equal to that of the baseline model when both share the same basic reproduction number ℛ_0_.

## 4 Discussion

Our analysis of COVID-19 mortality data from 30 US locations during the first pandemic wave reveals systematic biases that arise when epidemic models fail to account for behavioral responses to disease burden. Across all three behavioral models, at least 28 of 30 locations favored behavioral models—where transmission rates decline as mortality increases—over the traditional model with a constant transmission rate, based on both NSSE and Bayes factor. More critically, we identified a problematic paradox: models that ignore behavioral change systematically underestimate pathogen transmissibility while simultaneously overestimating total infection burden. We confirmed these biases through controlled synthetic experiments where the true dynamics were known by design, demonstrating that the discrepancies arise from model misspecification rather than data quality or stochastic effects. These findings held across three distinct mathematical formulations of behavioral models, strengthening confidence in our conclusions.

The dual bias we identify creates dangerous implications for public health decision-making. Models that ignore behavioral responses underestimate transmissibility as measured by the basic reproduction number, potentially delaying critical early interventions when aggressive action matters most. Yet these same models overestimate total infections, leading to excessive resource allocation for later epidemic phases that behavioral adaptation may prevent. Decision-makers relying on such models face a systematic mismatch: insufficiently aggressive early response combined with overpreparation for infection burdens that may not materialize. This paradox can result in both under-intervention during critical early phases and misallocation of resources for scenarios that behavioral change prevents.

The mechanism underlying these biases is intuitive once recognized. Baseline models with constant transmission rates fit observed mortality data using relatively modest transmission parameters because the transmission rate remains fixed throughout the entire epidemic wave. In contrast, behavioral models require substantially higher initial transmission rates to fit the same mortality data precisely because transmission declines as mortality increases and people adapt their behavior. This difference directly manifests in the basic reproduction number: the baseline model settles on a lower value that suffices under constant transmission, while the behavioral model correctly identifies the higher intrinsic transmissibility that characterized the pathogen before behavioral responses took effect. The bias in final epidemic size follows naturally from this same mechanism. When populations reduce contact rates in response to rising deaths—through actions like avoiding crowds, wearing masks, or increasing hygiene—fewer people ultimately become infected compared to scenarios where behavior remains unchanged.

This study has several important limitations. First, our models assume that behavioral changes are driven exclusively by location-specific reported mortality, whereas behavior in reality may also respond to mortality signals from nearby regions, national trends, policy announcements, or media coverage [6]. Second, using mortality as a proxy for public risk perception does not capture the complex array of influences on behavior, including policy actions, misinformation, informal information channels, and individual risk tolerance. This simplification may also introduce delays between changes in actual risk and behavioral adaptation, which our instantaneous feedback formulation does not capture [28, 29].

Third, we assume homogeneous populations within each location, ignoring substantial variation in risk tolerance, contact patterns, and protective behavior adoption across age groups, socioeconomic strata, and communities. Real populations exhibit heterogeneous responses to epidemic threats, with some groups adopting protective measures rapidly while others respond more slowly or not at all [6, 28–30]. Fourth, our analysis focuses exclusively on the first pandemic wave (March–July 2020), when behavioral responses were likely strongest due to novelty and uncertainty. Behavioral patterns may differ substantially in later waves as pandemic fatigue sets in or as populations develop more nuanced risk assessments [6].

Fifth, our models imply that extended post-peak mortality declines reflect stronger behavioral responses, but this relationship does not hold universally. New York experienced one of the earliest and most severe outbreaks [31–33] with stringent public health measures and well-documented behavioral changes [6, 34, 35], yet the mortality curve rose and fell relatively symmetrically without the prolonged tail our models associate with strong behavioral feedback. This disconnect suggests that the temporal signature of behavioral adaptation depends on factors we do not explicitly capture, such as the timing and stringency of policy interventions, underlying contact network structures, population density, or the rapidity with which protective behaviors were adopted. Locations with sharp mortality declines might reflect either weak behavioral responses or, conversely, such rapid behavioral adaptation that transmission was quickly suppressed. Our models cannot distinguish between these possibilities, which limits our ability to infer behavioral mechanisms from mortality data alone. Finally, our behavioral models assume a specific functional relationship between mortality and transmission reduction. While we tested three different functional forms, real behavioral responses may follow more complex patterns involving thresholds, saturation effects, or time-varying sensitivity to disease burden.

Future work should address these limitations through several avenues. Incorporating spatial coupling between locations could capture how mortality in nearby regions influences local behavior [36, 37]. Integrating data on policy interventions, mobility patterns, and direct behavioral surveys could provide more nuanced representations of behavioral change beyond mortality-driven responses. Age-structured models could account for heterogeneous contact patterns and risk perceptions across demographic groups [21]. Multi-wave analyses could reveal how behavioral responses evolve over the course of a pandemic as novelty effects diminish and risk perceptions stabilize. Comparing fits to multiple data streams—cases, hospitalizations, and deaths—could help disentangle behavioral responses to different epidemic indicators and their respective time lags.

More fundamentally, developing frameworks that explicitly model the cognitive and social processes underlying behavioral change—such as risk perception dynamics, information diffusion, and social norm formation— could provide deeper mechanistic understanding beyond phenomenological feedback functions [38]. Such frameworks could better predict behavioral responses to novel pathogens and inform more effective risk communication strategies.

Severing the coupling between pathogen transmission and human behavior produces models that systematically misrepresent reality. Infectious disease outbreaks unfold through feedback: transmission shapes behavior, and behavior alters transmission. Models that assume constant contact rates sacrifice this coupling for mathematical convenience, producing contradictory estimates of key epidemiological quantities— underestimating pathogen transmissibility while overestimating infection burden. These structural failures cascade through every decision that relies on model projections, from intervention timing to resource allocation.

Our findings demonstrate that incorporating behavioral dynamics is not merely a modeling refinement but essential for accurate epidemic parameter estimation and evidence-based pandemic response. Across 30 geographically and demographically diverse US locations, behavioral models consistently provided superior fits to observed data and more plausible estimates of both transmissibility and final epidemic size. The analytical results further establish that this dual bias is an inevitable mathematical consequence of ignoring behavioral feedback when it is present in the data. Future pandemic preparedness must prioritize the development and deployment of models that explicitly account for the dynamic interplay between disease transmission and human behavioral responses.

## Data Availability

The complete codebase used for simulation, inference, and figure generation is available at GitHub repository: https://github.com/markolalovic/epi-behavior-models

## Software and Data Availability

The complete codebase used for simulation, inference, and figure generation is available at GitHub repository: https://github.com/markolalovic/epi-behavior-models [26]. The COVID-19 mortality data used in this study are publicly available from the COVID-19 Data Repository maintained by the Center for Systems Science and Engineering (CSSE) at Johns Hopkins University: https://github.com/CSSEGISandData/COVID-19 [22, 23].

## Acknowledgments

BP and MS were supported by cooperative agreement CDC-RFA-FT-23-0069 from the CDC’s Center for Forecasting and Outbreak Analytics. The views expressed in written in this publication do not necessarily reflect the official policies of the Department of Health and Human Services/Centers for Disease Control and Prevention. ML acknowledges the PhD studentship support from Northeastern University London.

## A Proof of Theorem 3.1

*Proof*. Let *β*_*α*_(*t*) = *β*_0_ exp(−*αI*_*α*_(*t*)), where *α* = *ζδ* combines the behavioral sensitivity parameter *ζ* with the mortality rate *δ*, and *I*_*α*_(*t*) denotes the infectious population trajectory corresponding to *α*. Furthermore, we assume that the total population size is fixed with *N* = *S* + *E* + *I* + *R* + *D*.

The *final epidemic size* corresponding to *α* is defined as

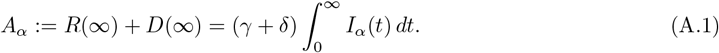

From the *S*–equation of model (2.1), we have

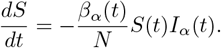

Dividing both sides by *S*(*t*) and integrating in time from 0 to ∞ (noting *S*(*t*) *>* 0 for all finite time until extinction):

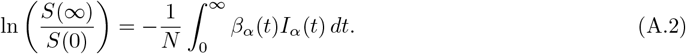

Since the removed individuals at infinity equal the initial population minus susceptibles at infinity, we have

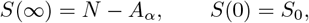

and hence, using *β*_*α*_(*t*) = *β*_0_ exp(−*αI*_*α*_(*t*)), (A.2) can be rewrriten as

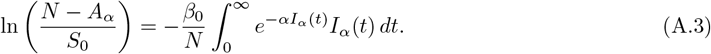

This holds for all *α* ≥ 0. When *α* = 0 (i.e. *β*(*t*) ≡ *β*_0_), we have

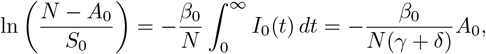

with *A*_0_ and *I*_0_(*t*) denoting the final size and infectious population corresponding to the baseline model. Hence, the classical implicit final-size equation:

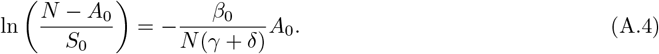

### Small-*α* expansion

We now expand for small *α >* 0. Let

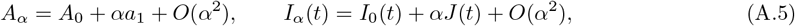

where, *a*_1_ and *J* (*t*) are the first-order corrections to the final epidemic size and infectious trajectory, respectively.

We expand both sides of (A.3) using the Taylor series.

### Left-hand side expansion

We expand ln 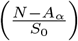 using Taylor series around *A* = *A*_0_:

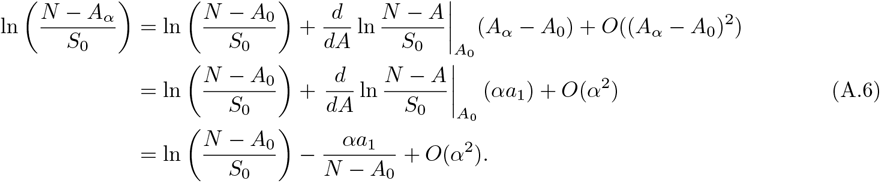

### Right-hand side expansion

Using 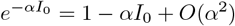 and *I*_*α*_ from (A.5), we expand:

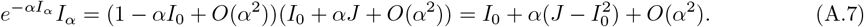

We expand the internal term in (A.3) using (A.7) as:

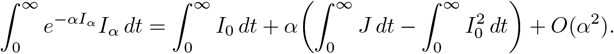

Plugging in the expansion of *A*_*α*_ and *I*_*α*_(*t*) from (A.5) into (A.1) and comparing *α* terms gives:

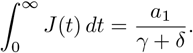

Hence, the right-hand side of (A.3) can be finally expanded as:

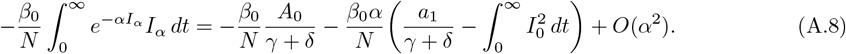

#### Matching terms

Now we compare the matching terms of left-hand side expansion, given in (A.6), with right-hand side expansion, given in (A.8). At order *O*(1), we recover the classical relation (A.4).

At order *O*(*α*) we have:

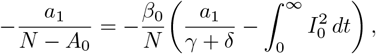

which can be rewritten as:

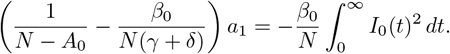

Therefore, solving for *a*_1_ gives:

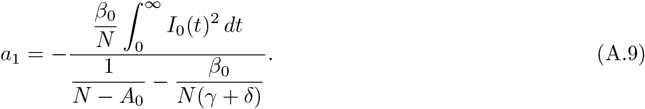

Since *A*_*α*_ = *A*_0_ + *αa*_1_ + *O*(*α*^2^), and *A*_0_ corresponds to the final size of the baseline model, to show that the final size of the behavioral model is smaller than that of the baseline model, it is sufficient to show that *a*_1_ *<* 0. Since we consider small *α*, the *O*(*α*^2^) terms are negligible compared to the first-order term *αa*_1_, so the sign of *a*_1_ determines whether *A*_*α*_ *< A*_0_ or *A*_*α*_ *> A*_0_.

The numerator of (A.9) is always positive. The denominator of (A.9) is positive provided:

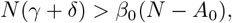

which can be rewritten as:

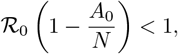

which is true since 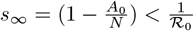 (see (B.9) for details). This concludes the proof.

## B Proof of Theorem 3.2

*Proof*. Let *β*_*α*_(*t*) = *β*_0_Ψ(*α, I*_*α*_(*t*)), where *α* = *ζδ* combines the behavioral sensitivity parameter *ζ* with the mortality rate *δ*, and *I*_*α*_(*t*) denotes the infectious population trajectory corresponding to *α*. For example, Ψ(*α, I*_*α*_(*t*)) = exp(−*ζδI*_*α*_(*t*)) = exp(−*αI*_*α*_(*t*)). We require that 0 *<* Ψ(*α, I*(*t*)) ≤ 1, and that Ψ(0, *I*(*t*)) = *β*_0_. We assume that the total population size is fixed with *N* = *S* +*E* +*I* +*R*+*D*. Furthermore, initial conditions are: *S*(0) = *S*_0_ *>* 0, *E*(0) = *E*_0_ ≥ 0, *I*(0) = *I*_0_ ≥ 0, *R*(0) = *D*(0) = 0 and *S*_0_ + *E*_0_ + *I*_0_ = *N* . The *final epidemic size* is defined as

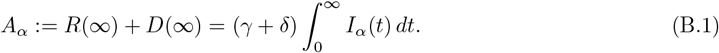

From the *S*–equation of model (2.1), we have

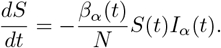

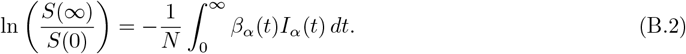

Since removed individuals at infinity equal the initial population minus susceptibles at infinity, we have

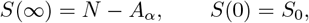

and hence (B.2) can be rewrriten as

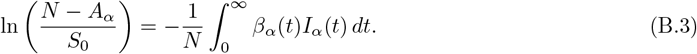

Equation (B.3) is exact for any time-dependent Ψ and any solution of the SEIRD model (2.1) satisfying the stated assumptions. Let *A*_0_ denote the final size when Ψ(*α* = 0, *I*(*t*)) = *β*_0_.

Recall the pointwise inequality assumption

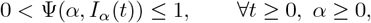

The above implies

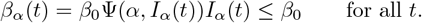

Substituting inequality of *β*_*α*_(*t*) in (B.3) gives:

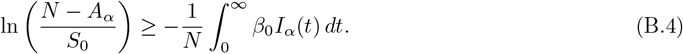

For the baseline case with *α* = 0 (no behavioral response), we have *β*_*α*_(*t*) = *β*_0_ and the infectious trajectory *I*_*α*_(*t*) reduces to *I*_0_(*t*). Substituting *β*_*α*_(*t*) = *β*_0_ in (B.3) yields:

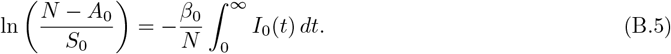

Equations (B.4) and (B.5) cannot be directly compared because they involve both different final epidemic sizes on the left hand side (*A*_*α*_ versus *A*_0_) and different infectious population trajectories on the right hand side (*I*_*α*_(*t*) versus *I*_0_(*t*)). To enable comparison, we apply the relation 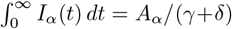 from equation (B.1), which transforms inequality (B.4) into:

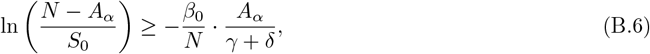

and for baseline case, equation (B.5) can rewritten as:

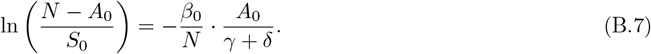

We define the function

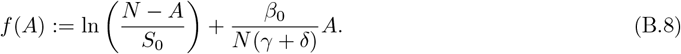

Notice that for the baseline model *f* (*A*_0_) = 0 while for behavioral models *f* (*A*_*α*_) ≥ 0. *f* (*A*) has two zeros — the trivial solution *A* = 0 (no epidemic) and the nontrivial solution *A* = *A*_0_ (baseline epidemic). The proof proceeds in three steps. First, we show *f* (*A*) is strictly concave and compute its unique maximum at *A*^∗^ = *N* (1 − 1*/*ℛ _0_). Second, since *A*_0_ is defined only implicitly through a transcendental equation (B.7), we cannot directly compare *A*_0_ with *A*^∗^. We therefore normalize the susceptible population as *s* = *S/N* and express the baseline final size through the transcendental equation ln *s*_∞_ = −ℛ_0_(1 − *s*_∞_), where *s*_∞_ = (*N* − *A*_0_)*/N* . By analyzing the auxiliary function Φ(*s*) := ln *s* + ℛ_0_(1 − *s*), we show that it attains a unique maximum at *s* = 1*/*ℛ_0_. Furthermore, we prove that *s*_∞_ *<* 1*/*ℛ_0_, which translates to *A*_0_ *> A*^∗^. Third, combining *A*_0_ *> A*^∗^ with the concavity of *f* (*A*) and the inequality *f* (*A*_*α*_) ≥ 0, we conclude *A*_*α*_ ≤ *A*_0_. Details follow.

Differentiating *f* (*A*) with respect to *A* gives (where 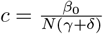 is a constant):

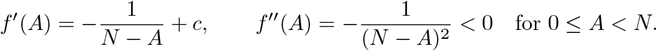

Thus *f* is strictly concave on [0, *N*) and can have at most one critical point (which will be a global maximum).

Solving 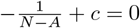, which gives 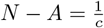. Thus the unique critical point is

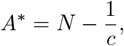

where *c* = ℛ_0_*/N* . Because *f* ^*′′*^(*A*) *<* 0, *A*^∗^ is the global maximiser of *f* on [0, *N*).

### Relation to *A*_0_

Introduce the normalized susceptible fraction *s* := *S/N*, and assume that *S*_0_ = *N* . It follows from (B.7) that the classical final-size equation for *α* = 0 can be written in normalized form as

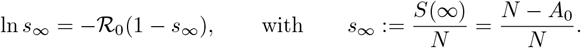

Define the auxiliary function

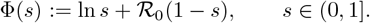

Then *s*_∞_ is a zero of Φ, i.e. Φ(*s*_∞_) = 0.

Compute the derivative:

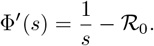

Hence Φ^*′*^(*s*) *>* 0 for 0 *< s <* 1*/*ℛ_0_, Φ^*′*^(1*/*ℛ_0_) = 0, and Φ^*′*^(*s*) *<* 0 for *s >* 1*/*ℛ_0_. Thus Φ attains its unique maximum at *s* = 1*/*ℛ_0_.

Evaluate Φ at the endpoints and at the maximum:

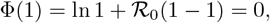

and by the standard inequality ln *x* ≤ *x* − 1 (with equality only at *x* = 1),

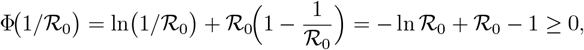

with strict inequality when ℛ_0_ = 1. Thus the maximum value Φ(1*/*ℛ_0_) is nonnegative (positive for ℛ_0_ *>* 1) while Φ(1) = 0.

Because Φ is continuous on (0, 1], has a unique maximum at 1*/*ℛ_0_ with Φ(1*/*ℛ_0_) ≥ 0, and Φ(1) = 0, the equation Φ(*s*) = 0 has exactly two solutions in (0, 1] when Φ(1*/*ℛ_0_) *>* 0: one at *s* = 1 and the other strictly to the left of 1*/*ℛ_0_. The epidemiologically relevant solution is the nontrivial root *s*_∞_ ∈ (0, 1). Because Φ(1*/*ℛ_0_) *>* 0 and Φ(1) = 0, this nontrivial root must satisfy

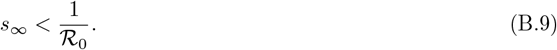

### Conclude *A*^∗^ *< A*_0_

Translate back to *A*. Since *s*_∞_ = (*N* − *A*_0_)*/N*, the inequality *s*_∞_ *<* 1*/*ℛ_0_ is equivalent to

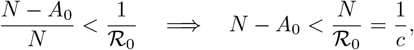

Hence

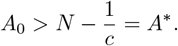

Therefore the unique maximiser *A*^∗^ of *f* (*A*), given by (B.8), indeed lies strictly to the left of the classical final size *A*_0_ (for ℛ_0_ *>* 1, i.e. in the nontrivial epidemic regime). Since *A*_*α*_ satisfies inequality (B.6), we conclude that *A*_*α*_ must lie to the left of *A*_0_. Hence, *A*_*α*_ ≤ *A*_0_.

