## Supplementary material for "The Paradox of Neglecting Changes in Behavior: How Standard Epidemic Models Misestimate Both Transmissibility and Final Epidemic Size": supp

### Contents

**S1 Excluded U.S. States** **3**

**S2 Parameter Inference Details** **4**

**S3 Synthetic Data Experiment Details** **5**

**S4 Exponential and Rational Behavioral Models** **8**

**S5 Sensitivity on Zeta** **14**

**S6 Effective Transmission Rate** **15**

**S7 Effective Reproduction Number** **18**

### List of Tables

S1 Comparison of the baseline model and the behavioral (Exponential) model across 30 U.S. states. 12
S2 Comparison of the baseline model and the behavioral (Rational) model across 30 U.S. states. 13

### S1 Excluded U.S. States

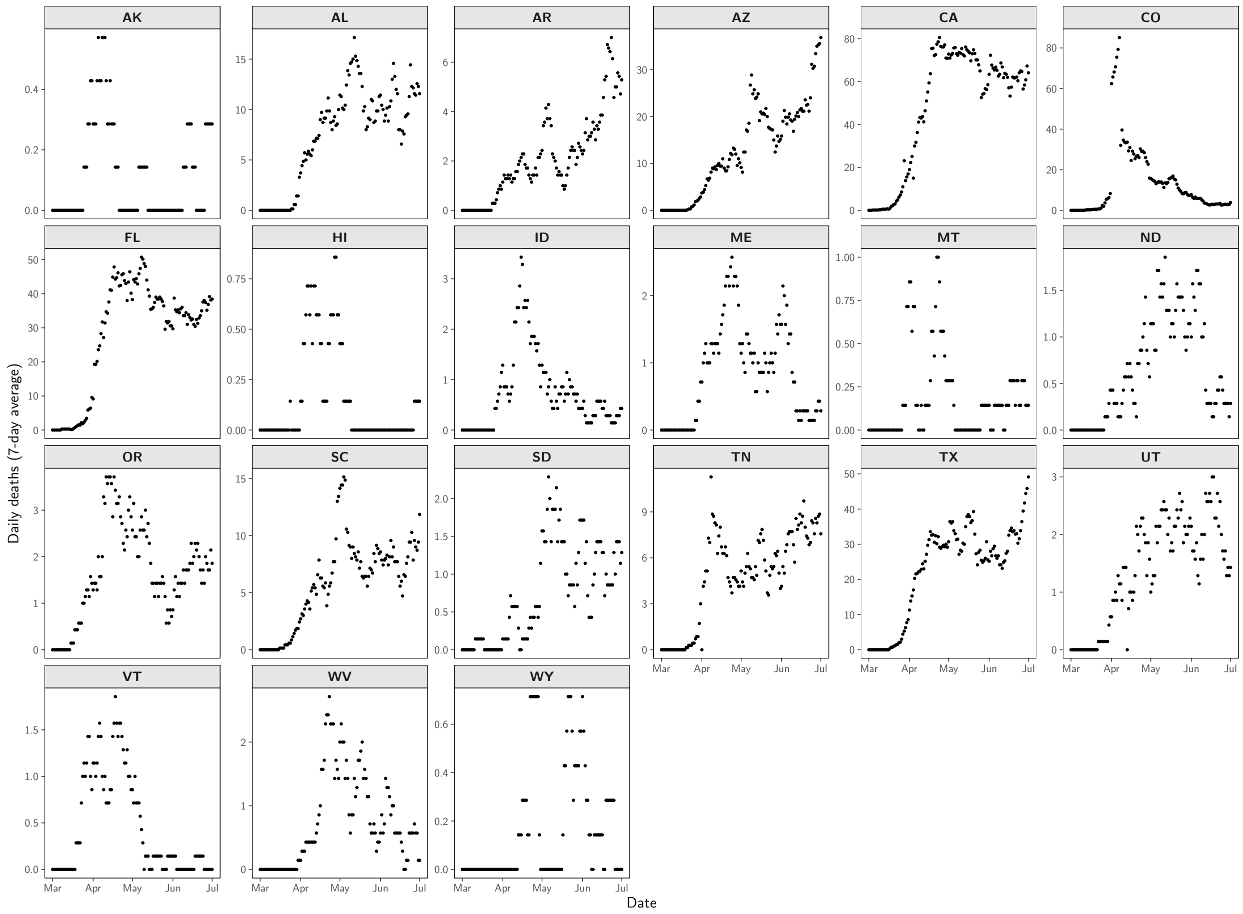

**Figure S1: COVID-19 mortality time series for U.S. states excluded from the main analysis.** Panels show observed 7-day averaged daily COVID-19 deaths for the 21 U.S. states ordered alphabetically that did not meet our inclusion criteria. States were excluded due to (1) very low daily death counts, (2) absence of a clear single-wave structure within the observation window, or (3) apparent reporting anomalies such as discontinuities in cumulative deaths, see Methods: Data and State Selection 2.2.

### S2 Parameter Inference Details

This section provides additional implementation details that complement the parameter inference description in the main text (Section 2.3), without repeating the full specification of the model, priors, or ABC–SMC setup.

**Data and preprocessing.** For each state, we work with state-level COVID-19 mortality over the period of $T = 123$ days from March 1 to July 1, 2020. Let $D_{\text{obs}}(t)$ denote cumulative reported deaths. Observed daily mortality counts are obtained as

$$M_{\text{obs}}(t) = D_{\text{obs}}(t) - D_{\text{obs}}(t-1). \tag{S2.1}$$

To suppress reporting artifacts such as weekend backlogs, we apply a trailing 7-day moving average,

$$\widetilde{M}(t) = \frac{1}{7}\sum_{k=0}^{6} M(t-k). \tag{S2.2}$$

Smoothing is performed over an extended window starting six days before the analysis start date. The first six padded values are discarded so that the final smoothed series $\big(\widetilde{M}(t)\big)_{t=1}^{T}$ covers exactly the target interval.

The same preprocessing is applied to model-simulated trajectories. For each simulation, the SEIRD system (Eq. (2.1)) is integrated for $T + 6$ days, daily deaths are computed as in Eq. (S2.1), smoothed using the trailing 7-day moving average (S2.2), and then truncated to the same $T$-day window as the data. Because identical smoothing is applied to both observed and simulated series, the small delay introduced by the moving average cancels in the distance calculation.

**Fixed parameters.** Fixed parameters are chosen based on early SARS-CoV-2 data. The incubation rate was set to $\sigma = 1/3$ day$^{-1}$ (average latent period of 3 days) [1, 2], and the recovery rate to $\gamma = 1/10$ day$^{-1}$ (average infectious period of 10 days) [3, 4]. The disease-induced mortality rate $\delta$ is inferred rather than fixed.

The total population size $N$ is location-specific and obtained from the processed Johns Hopkins CSSE data. Initial conditions are fixed at $E_0 = 1$, $R_0 = 0$, and $D_0 = 0$, with $I_0$ determined from the inferred initial prevalence and $S_0 = N - (E_0 + I_0 + R_0)$.

**Inferred parameters and reparameterization.** As described in the main text, we reparameterize the model in terms of the basic reproduction number $\mathcal{R}_0$ and the initial prevalence $\pi_0 = I_0/N$ rather than directly estimating $\beta_0$ and $I_0$. At each evaluation,

$$\beta_0 = \mathcal{R}_0(\gamma + \delta), \qquad I_0 = \max\{1,\, N\pi_0\},$$

so that $\mathcal{R}_0$ retains its standard interpretation as the basic reproduction number in a fully susceptible population (Eq. (2.6)).

The priors on $\mathcal{R}_0, \pi_0, \delta, \zeta$ are given in Section 2.3. Here we simply note that log-uniform (Jeffreys-type) priors on $\pi_0$ and $\delta$ are used to allow these positive parameters to vary over several orders of magnitude without favoring any particular scale a priori, which is appropriate when both are expected to be small and poorly constrained before observing the data.

**Parameter estimation via ABC–SMC.** We compute the discrepancy between simulated $\boldsymbol{y}_{\text{sim}}(\boldsymbol{\theta}) = \big(\widetilde{M}_{\text{sim}}(t;\boldsymbol{\theta})\big)_{t=1}^{T}$ and observed smoothed mortality series $\boldsymbol{y}_{\text{obs}} = \big(\widetilde{M}_{\text{obs}}(t)\big)_{t=1}^{T}$ using the normalized sum of squared errors (NSSE) defined in Eq. (2.9). Each ABC–SMC run uses $P = 1000$ particles per generation, a multivariate normal transition kernel with covariance adapted from the previous population, and a quantile tolerance schedule with $\alpha = 0.3$ for up to 10 generations: the tolerance at generation $t + 1$ is set to the $\alpha$-quantile of the accepted distances at generation $t$.

**Quantile tolerance schedule.** At each generation $t$ the tolerance $\varepsilon_t$ determines the maximum allowable discrepancy for accepting a particle. Rather than fixing $\varepsilon_t$ in advance, we adapt it using a quantile schedule: $\varepsilon_{t+1}$ is set to the 30th percentile of the distances of the accepted particles at generation $t$. This adaptive schedule gradually tightens the tolerance, focusing sampling on regions of higher posterior density while maintaining reasonable acceptance rates and avoiding premature loss of particle diversity.

**Fairness of calibration across models and locations.** The same ABC–SMC configuration (particle count, tolerance schedule, transition kernel, stopping criteria) is used for the baseline model and all behavioral variants across all locations to ensure a fair comparison. No model or location received preferential tolerances, evaluation budgets, or tuning. Consequently, differences in fit quality, inferred parameters, and model probabilities reflect model structure and data characteristics rather than differences in calibration effort.

### S3 Synthetic Data Experiment Details

The main text describes the design of the synthetic data experiments: mortality trajectories are generated from behavioral models with varying behavioral sensitivity $\zeta$, small Gaussian noise is added, and both baseline and behavioral models are fitted to these synthetic datasets to assess bias and identifiability. Here we provide implementation details not stated in the main text.

**Data-generating parameters.** Synthetic mortality trajectories were generated using the behavioral (Mixed) model (Eq. (2.4)) with parameters calibrated to Massachusetts COVID-19 mortality data. The total population size $N$ was fixed to the Massachusetts population, and the parameters $\pi_0$, $\mathcal{R}_0$, and $\delta$ were set to the weighted medians of their posterior marginal distributions under the behavioral (Mixed) model for Massachusetts. The behavioral sensitivity parameter $\zeta$ was varied across six values to represent increasing feedback strength:

$$\zeta \in \{0.001, 0.005, 0.009, 0.012, 0.016, 0.02\}.$$

Each synthetic dataset (for a given $\zeta$ and behavioral formulation) was treated as an observed time series and analyzed using the same ABC–SMC configuration as in the real-data analysis (see Section 2.3).

The main text focuses on the behavioral (Mixed) formulation (Figure 3). Supplementary Figures S2 and S3 show the corresponding results when the data-generating model uses the exponential and rational formulations, respectively.

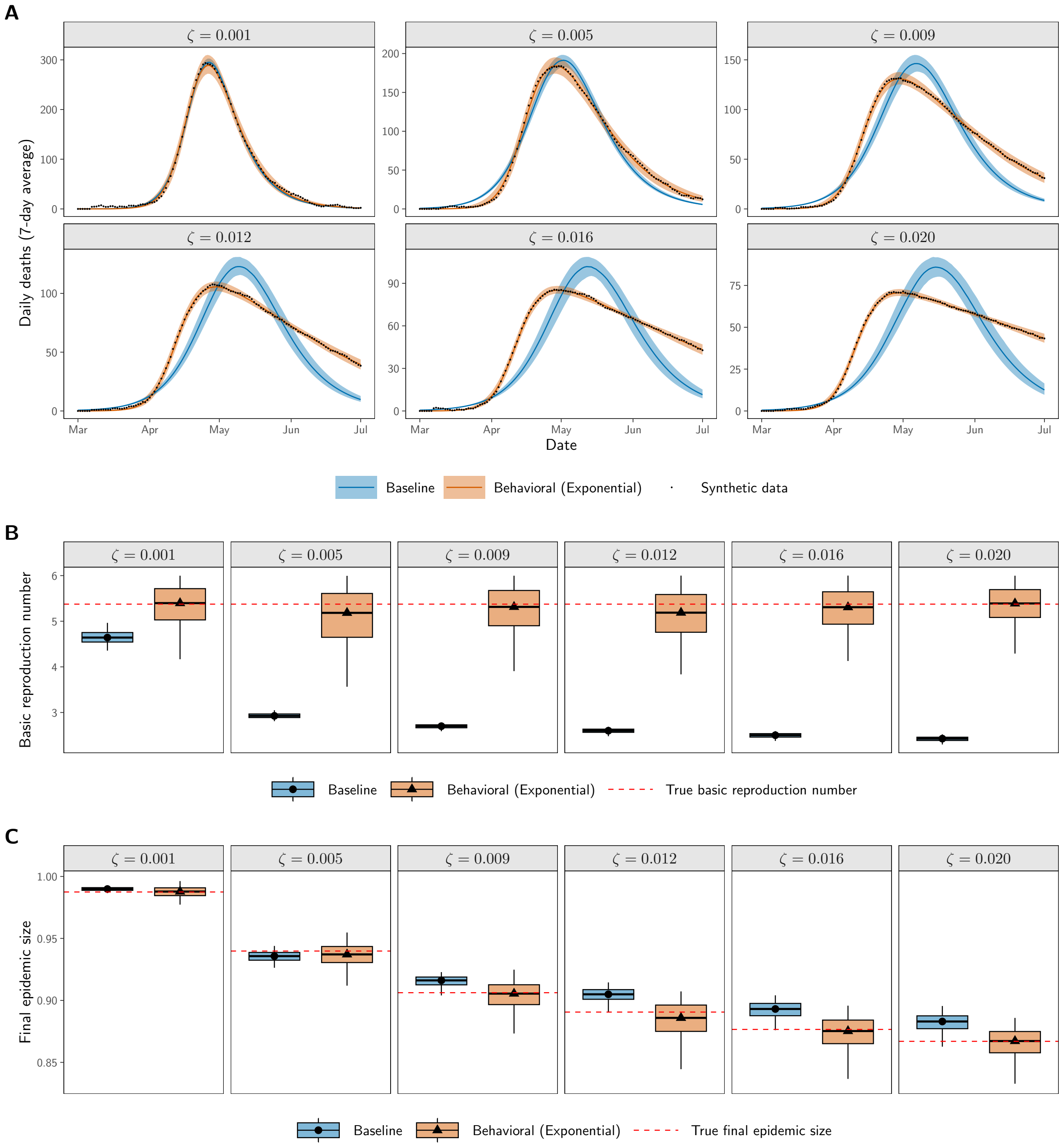

**Figure S2: Synthetic data experiment comparing baseline and behavioral (Exponential) models.** Synthetic data experiments where mortality trajectories were generated using the behavioral (Exponential) model with known parameters and varying behavioral sensitivity $\zeta$, then fitted with both baseline and behavioral (Exponential) models. **(A)** Posterior predictive fits to synthetic mortality data (black points). Solid lines show posterior medians and shaded regions show 90% credible intervals. The behavioral model accurately recovers the true dynamics across all $\zeta$ values, while the baseline model increasingly fails to capture the extended post-peak decline as behavioral feedback strengthens. **(B)** Posterior distributions of the basic reproduction number $\mathcal{R}_0$. The behavioral model recovers the true $\mathcal{R}_0$ (red dashed line) across all scenarios, while the baseline model systematically underestimates $\mathcal{R}_0$, with bias increasing as $\zeta$ increases. **(C)** Posterior distributions of the final epidemic size.

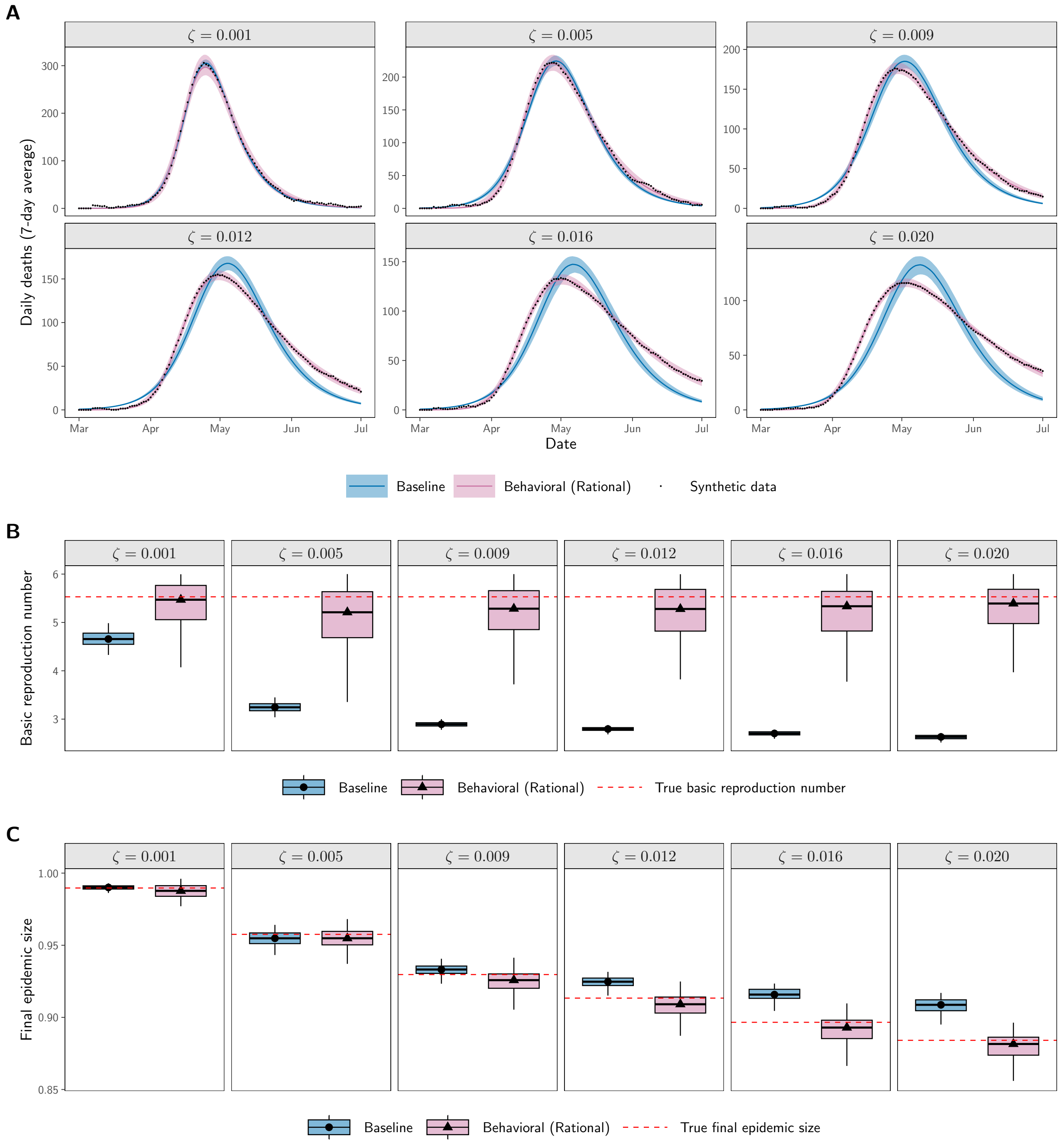

**Figure S3: Synthetic data experiment comparing baseline and behavioral (Rational) models.** Synthetic data experiments where mortality trajectories were generated using the behavioral (Rational) model with known parameters and varying behavioral sensitivity $\zeta$, then fitted with both baseline and behavioral (Rational) models. **(A)** Posterior predictive fits to synthetic mortality data (black points). Solid lines show posterior medians and shaded regions show 90% credible intervals. The behavioral model accurately recovers the true dynamics across all $\zeta$ values, while the baseline model increasingly fails to capture the extended post-peak decline as behavioral feedback strengthens. **(B)** Posterior distributions of the basic reproduction number $\mathcal{R}_0$. The behavioral model recovers the true $\mathcal{R}_0$ (red dashed line) across all scenarios, while the baseline model systematically underestimates $\mathcal{R}_0$, with bias increasing as $\zeta$ increases. **(C)** Posterior distributions of the final epidemic size.

### S4 Exponential and Rational Behavioral Models

The main text focuses on the behavioral (Mixed) model when presenting posterior predictive fits and comparisons of $\mathcal{R}_0$ and final epidemic size (Figures 1 and 2). Here we provide corresponding summaries for the exponential and rational behavioral formulations.

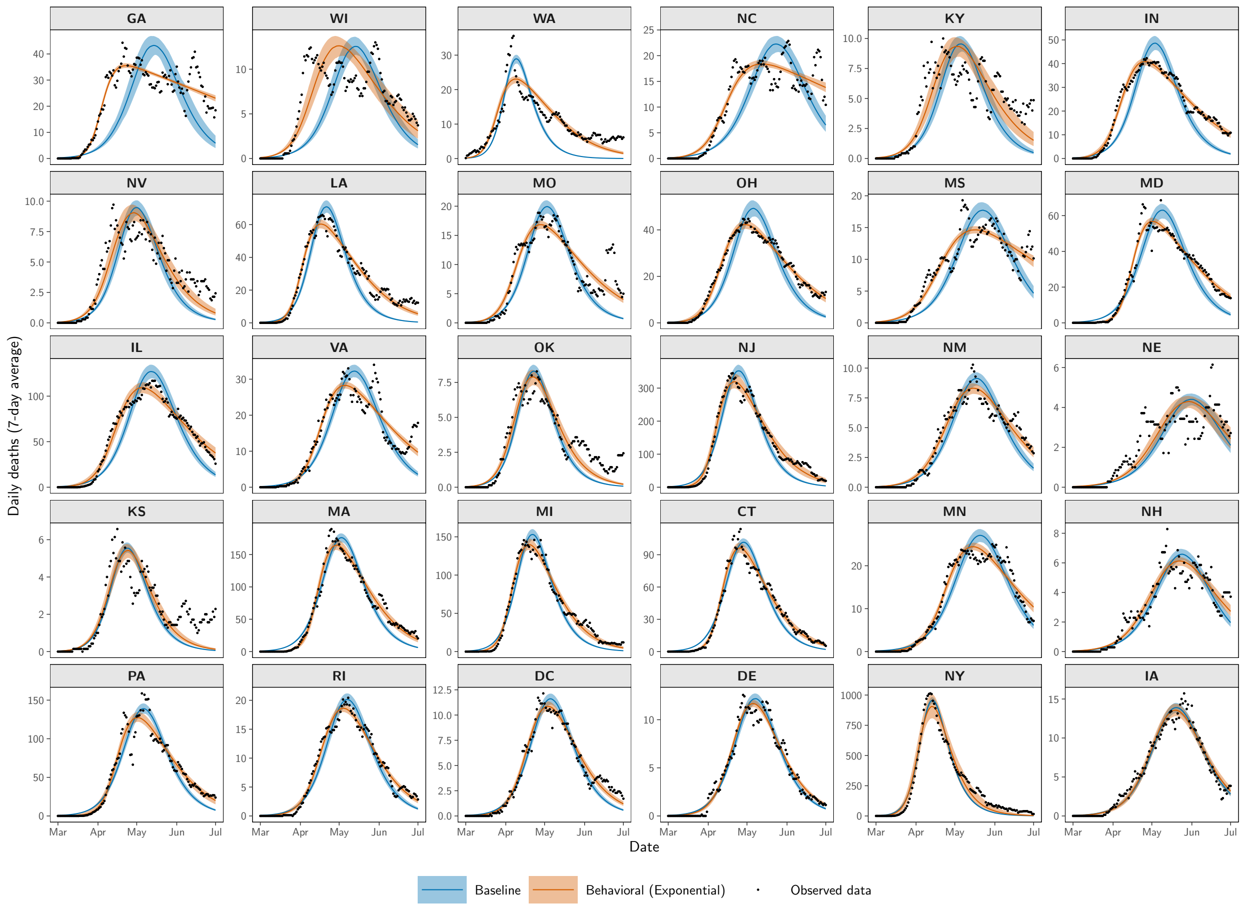

**Figure S4: Behavioral (Exponential) model fits compared to baseline across 30 U.S. locations.** Posterior predictive fits comparing baseline and behavioral (Exponential) models to 7-day averaged daily COVID-19 deaths during the first pandemic wave (March–July 2020). Black points show observed mortality data. Solid lines show posterior predictive medians and shaded regions show 90% credible intervals (5th–95th percentiles), computed from 1,000 weighted posterior samples from the final ABC–SMC generation. The behavioral (Exponential) model captures the characteristic asymmetric mortality curves with sharp initial rises and gradual extended declines observed in many locations, whereas the baseline model with constant transmission often decays too rapidly. States are ordered from left to right and top to bottom by decreasing improvement in fit, measured by the difference in median posterior NSSE between the behavioral and baseline models, analogous to Figure 1 in the main text.

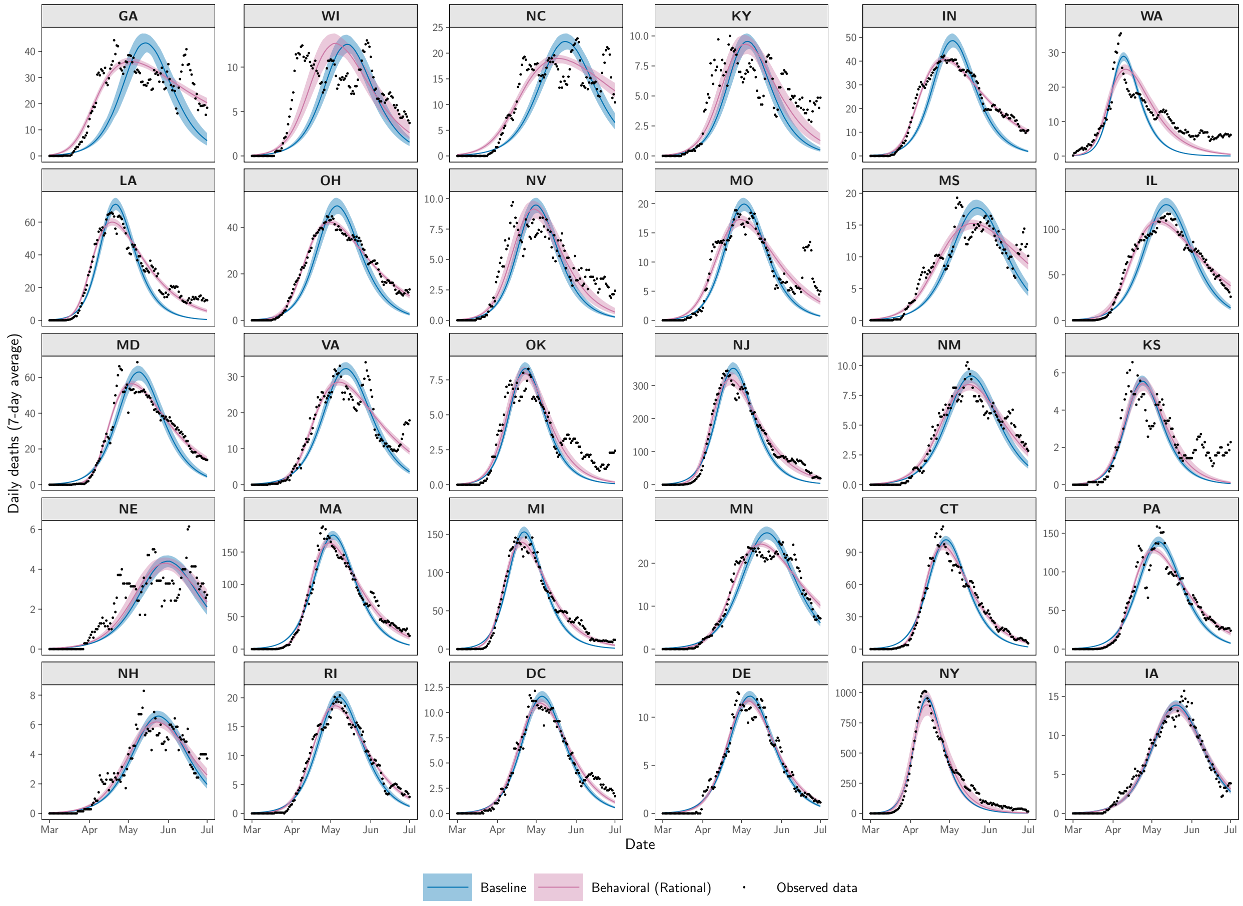

**Figure S5: Behavioral (Rational) model fits compared to baseline across 30 U.S. locations.** Posterior predictive fits comparing baseline and behavioral (Rational) models to 7-day averaged daily COVID-19 deaths during the first pandemic wave (March–July 2020). Black points show observed mortality data. Solid lines show posterior predictive medians and shaded regions show 90% credible intervals (5th–95th percentiles), computed from 1,000 weighted posterior samples from the final ABC–SMC generation. The behavioral (Rational) model also reproduces the asymmetric mortality curves with sharp rises and extended declines that are difficult to match under a constant-transmission baseline model. States are ordered from left to right and top to bottom by decreasing improvement in fit, measured by the difference in median posterior NSSE between the behavioral and baseline models, analogous to Figure 1 in the main text.

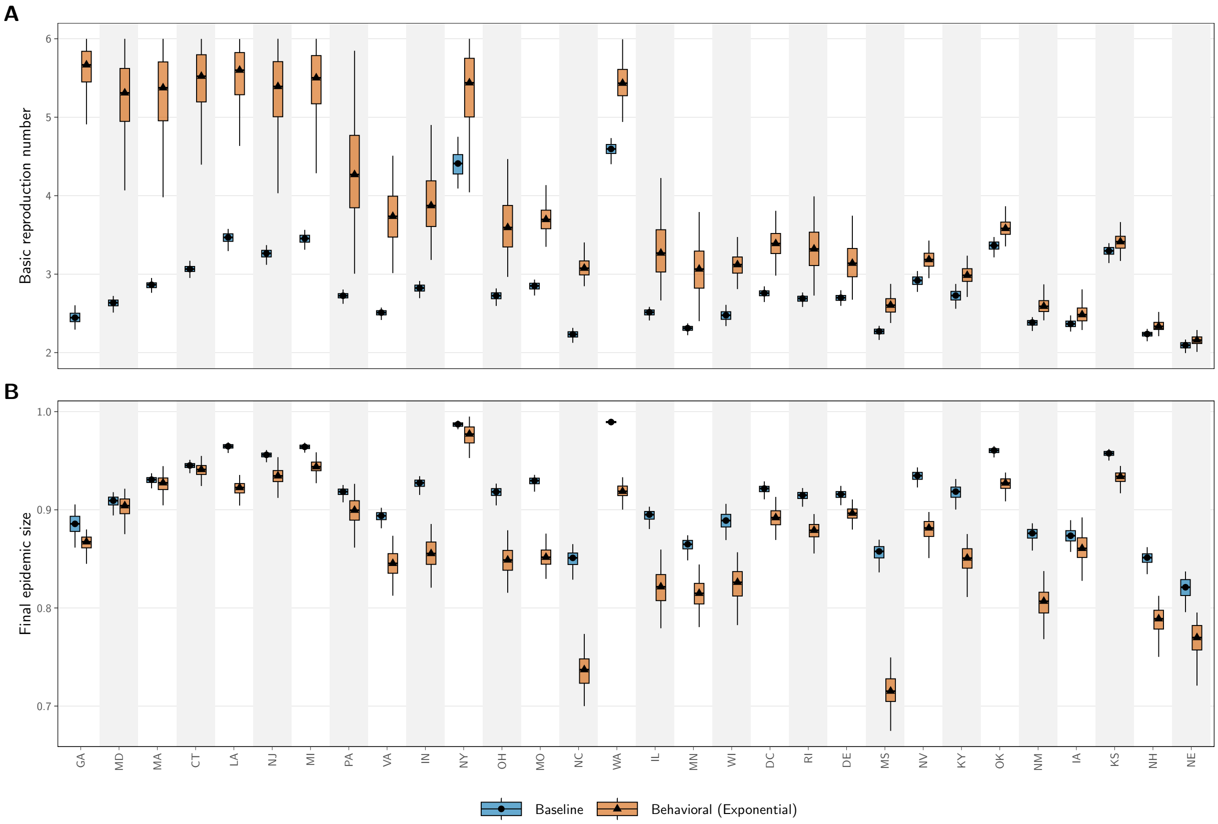

**Figure S6: Baseline versus behavioral (Exponential) model estimates of $\mathcal{R}_0$ and final epidemic size.** Posterior distributions of the basic reproduction number $\mathcal{R}_0$ and final epidemic size for the baseline and behavioral (Exponential) models fitted to COVID-19 mortality data from 30 U.S. locations during the first pandemic wave (March–July 2020). Boxplots show median, interquartile range, and whiskers extending to $1.5 \times \text{IQR}$ within the support of weighted posterior samples. States are ordered by decreasing difference in median $\mathcal{R}_0$ between the behavioral (Exponential) and baseline models.

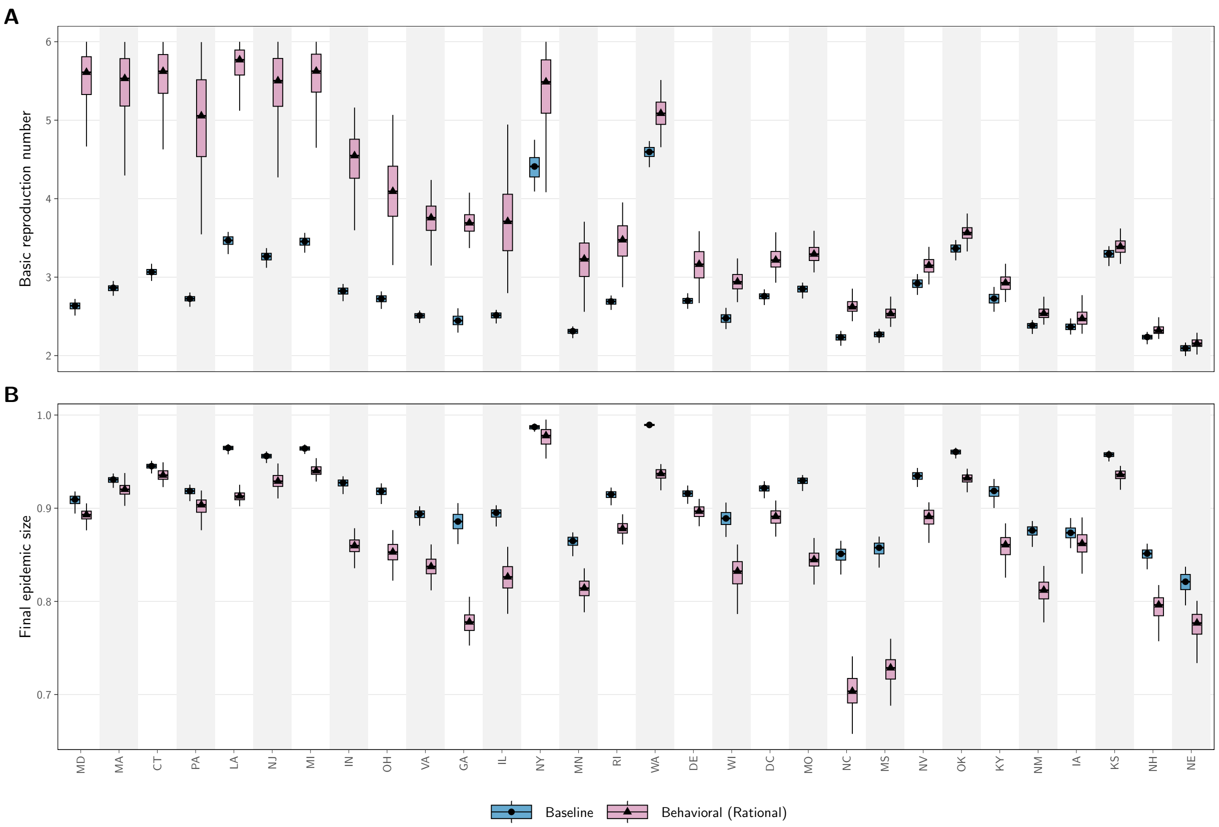

**Figure S7: Baseline versus behavioral (Rational) model estimates of $\mathcal{R}_0$ and final epidemic size.** Posterior distributions of the basic reproduction number $\mathcal{R}_0$ and final epidemic size for the baseline and behavioral (Rational) models fitted to COVID-19 mortality data from 30 U.S. locations during the first pandemic wave (March–July 2020). Boxplots show median, interquartile range, and whiskers extending to $1.5 \times$ IQR within the support of weighted posterior samples. States are ordered by decreasing difference in median $\mathcal{R}_0$ between the behavioral (Rational) and baseline models.

**Table S1:** Comparison of the baseline model and the behavioral (Exponential) model across 30 U.S. states. For each state, Median $NSSE_B$ and Median $NSSE_E$ denote the median posterior normalized sum of squared errors (NSSE) under the baseline $B$ and behavioral (Exponential) $E$ models respectively. $BF_{eq}$ is the Bayes factor under equal model priors in favor of the exponential model versus the baseline model at the stabilized ABC–SMC generation, and "Evidence" gives the corresponding verbal strength of evidence.

| State | Median $NSSE_B$ | Median $NSSE_E$ | $BF_{eq}$ | Evidence |
|---|---|---|---|---|
| NH | 0.0571 | **0.0401** | 20035.30 | very strong |
| VA | 0.0805 | **0.0385** | 9343.72 | very strong |
| NC | 0.1440 | **0.0363** | 3660.24 | very strong |
| MO | 0.1214 | **0.0573** | 2326.63 | very strong |
| RI | 0.0250 | **0.0105** | 529.25 | very strong |
| GA | 0.2254 | **0.0260** | 577.91 | very strong |
| WI | 0.2540 | **0.1139** | 335.87 | very strong |
| OH | 0.0690 | **0.0054** | 190.33 | very strong |
| MS | 0.0882 | **0.0338** | 133.93 | strong |
| NM | 0.0488 | **0.0195** | 124.12 | strong |
| IN | 0.0896 | **0.0087** | 104.37 | strong |
| CT | 0.0330 | **0.0110** | 69.45 | strong |
| IL | 0.0572 | **0.0110** | 61.22 | strong |
| MN | 0.0392 | **0.0184** | 59.66 | strong |
| KY | 0.2154 | **0.1128** | 31.04 | strong |
| MD | 0.0621 | **0.0144** | 26.79 | strong |
| PA | 0.0471 | **0.0311** | 20.64 | strong |
| NJ | 0.0433 | **0.0130** | 18.37 | positive |
| MA | 0.0374 | **0.0131** | 16.73 | positive |
| DE | 0.0210 | **0.0162** | 16.53 | positive |
| LA | 0.0842 | **0.0161** | 16.33 | positive |
| MI | 0.0344 | **0.0109** | 8.05 | positive |
| WA | 0.1786 | **0.0681** | 8.05 | positive |
| NE | 0.1390 | **0.1104** | 1.27 | very weak |
| DC | 0.0285 | **0.0147** | 1.17 | very weak |
| NV | 0.1456 | **0.0734** | 1.17 | very weak |
| OK | 0.1005 | **0.0629** | 1.17 | very weak |
| KS | 0.1627 | **0.1341** | 1.06 | very weak |
| IA | **0.0141** | 0.0165 | 0.79 | favors Baseline |
| NY | 0.0277 | **0.0273** | 0.03 | favors Baseline |

**Table S2:** Comparison of the baseline model and the behavioral (Rational) model across 30 U.S. states. For each state, Median $NSSE_B$ and Median $NSSE_R$ denote the median posterior normalized sum of squared errors (NSSE) under the baseline $B$ and behavioral (Rational) $R$ models respectively. $BF_{eq}$ is the Bayes factor under equal model priors in favor of the rational model versus the baseline model at the stabilized ABC–SMC generation, and "Evidence" gives the corresponding verbal strength of evidence.

| State | Median $NSSE_B$ | Median $NSSE_R$ | $BF_{eq}$ | Evidence |
|---|---|---|---|---|
| GA | 0.2254 | **0.0363** | 21622.10 | very strong |
| RI | 0.0250 | **0.0098** | 580.79 | very strong |
| LA | 0.0842 | **0.0169** | 484.47 | very strong |
| VA | 0.0805 | **0.0384** | 551.33 | very strong |
| MI | 0.0344 | **0.0113** | 618.29 | very strong |
| MN | 0.0392 | **0.0176** | 766.21 | very strong |
| NE | 0.1390 | **0.1135** | 684.73 | very strong |
| MO | 0.1214 | **0.0634** | 378.67 | very strong |
| WI | 0.2540 | **0.1432** | 235.46 | very strong |
| MD | 0.0621 | **0.0154** | 260.53 | very strong |
| OH | 0.0690 | **0.0044** | 75.25 | strong |
| IL | 0.0572 | **0.0088** | 147.28 | strong |
| IN | 0.0896 | **0.0072** | 99.39 | strong |
| MS | 0.0882 | **0.0352** | 114.97 | strong |
| NC | 0.1440 | **0.0449** | 113.90 | strong |
| KY | 0.2154 | **0.1299** | 48.24 | strong |
| PA | 0.0471 | **0.0290** | 49.68 | strong |
| NV | 0.1456 | **0.0859** | 38.68 | strong |
| OK | 0.1005 | **0.0681** | 36.42 | strong |
| MA | 0.0374 | **0.0140** | 23.95 | strong |
| NJ | 0.0433 | **0.0134** | 22.78 | strong |
| NM | 0.0488 | **0.0215** | 21.02 | strong |
| WA | 0.1786 | **0.0997** | 20.84 | strong |
| DE | 0.0210 | **0.0160** | 15.75 | positive |
| KS | 0.1627 | **0.1372** | 12.89 | positive |
| IA | **0.0141** | 0.0158 | 2.37 | very weak |
| CT | 0.0330 | **0.0123** | 1.76 | very weak |
| NH | 0.0571 | **0.0413** | 1.24 | very weak |
| DC | 0.0285 | **0.0156** | 1.15 | very weak |
| NY | **0.0277** | 0.0285 | 0.04 | favors Baseline |

#### S5 Sensitivity on Zeta

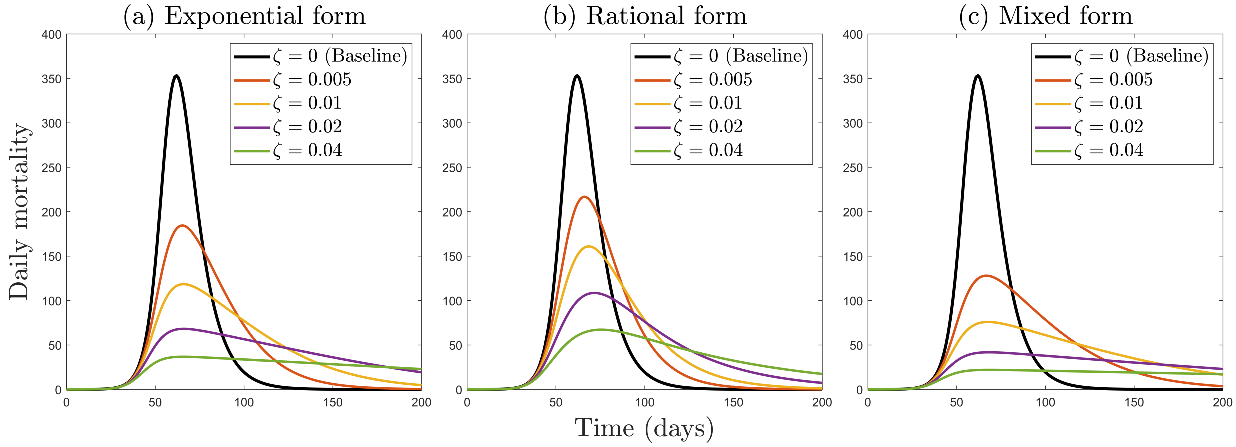

**Figure S8:** Daily mortality curves for three behavioral models with varying behavioral response strength $\zeta$. (a) Exponential form: $\beta(t) = \beta_0 e^{-\zeta\delta I(t)}$, where transmission rate decays exponentially with increasing infections. (b) Rational form: $\beta(t) = \beta_0/(1 + \zeta\delta I(t))$, which saturates more slowly as infections rise. (c) Mixed form: $\beta(t) = \beta_0 e^{-\zeta\delta I(t)}/(1 + \zeta\delta I(t))$, combining both exponential and rational responses. The baseline model ($\zeta = 0$, black line) represents traditional SEIR dynamics without behavioral adaptation. Stronger behavioral responses (higher $\zeta$ values) lead to lower peak mortality and more prolonged epidemic tails. Model parameters: $\sigma = 1/3$ day$^{-1}$, $\gamma = 1/10$ day$^{-1}$, $\delta = 0.001$, $\beta_0 = 0.5$, $N = 10^6$.

#### S6 Effective Transmission Rate

For each model variant and state, we computed the posterior distribution of the effective transmission rate $\beta(t)$ by propagating uncertainty from the ABC–SMC parameter posteriors. From each state- and model-specific posterior, we drew 1,000 parameter sets, where the probability of selecting a posterior particle is proportional to its ABC–SMC importance weight. For each sampled parameter set we simulated the SEIRD model, extracted the aligned infectious trajectory $I(t)$, and computed the corresponding effective transmission rate

$$\beta_{\text{eff}}(t) = \begin{cases} \beta_0, & \text{Baseline}, \\ \beta_0\, e^{-\zeta\,\delta\,I(t)}, & \text{Behavioral (Exponential)}, \\ \frac{\beta_0}{1+\zeta\,\delta\,I(t)}, & \text{Behavioral (Rational)}, \\ \frac{\beta_0\, e^{-\zeta\,\delta\,I(t)}}{1+\zeta\,\delta\,I(t)}, & \text{Behavioral (Mixed)}. \end{cases}$$

Pointwise 5th, 50th, and 95th percentiles across the ensemble of 1,000 trajectories form the median curve and the 90% pointwise credible interval bands shown in Figures S9, S10, and S11.

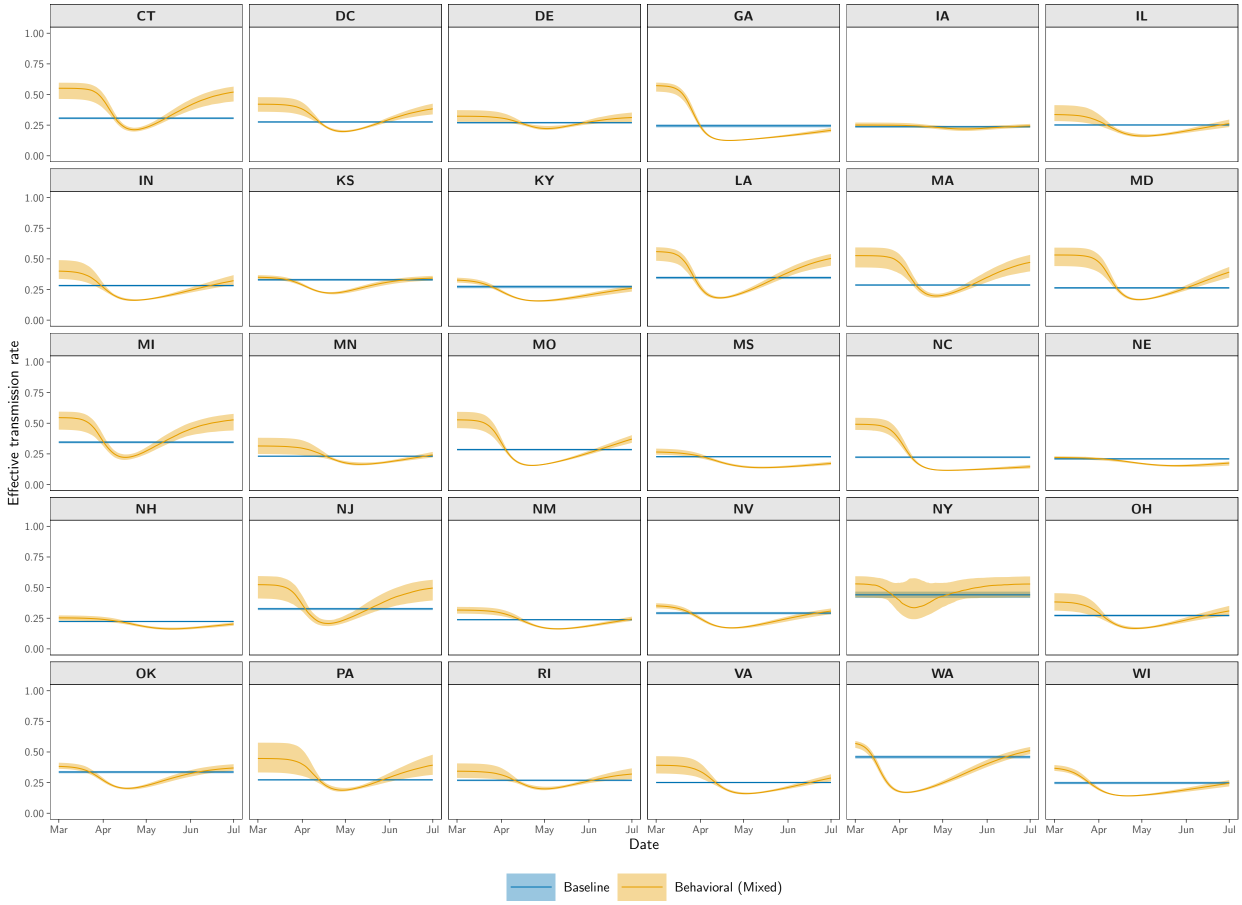

**Figure S9: Effective transmission rate $\beta(t)$ under the Behavioral (Mixed) model.** Shown are posterior median trajectories together with 90% pointwise credible intervals, obtained by propagating weighted ABC–SMC posterior samples through the SEIRD model to compute $\beta(t)$ for each draw.

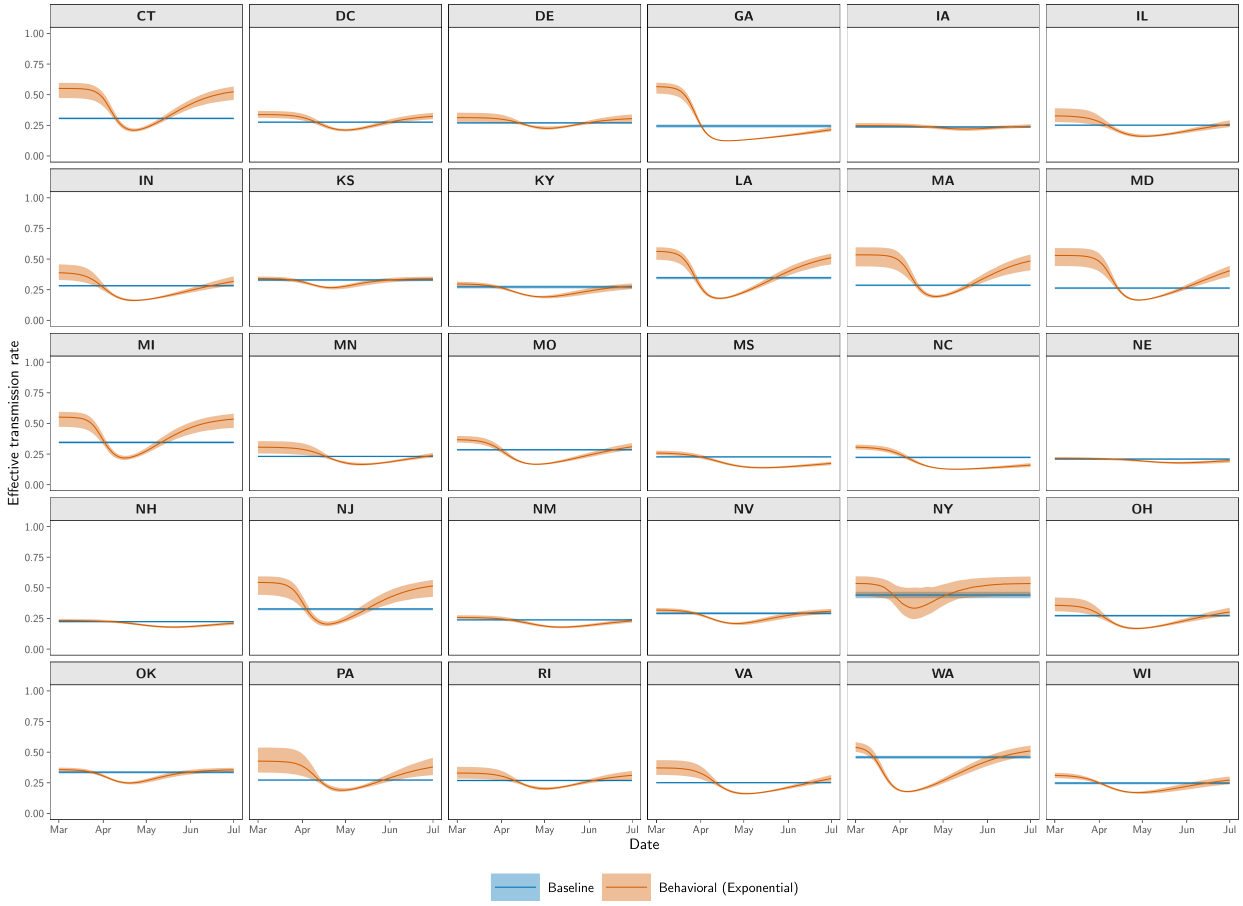

**Figure S10: Effective transmission rate $\beta(t)$ under the Behavioral (Exponential) model.** Shown are posterior median trajectories together with 90% pointwise credible intervals, obtained by propagating weighted ABC–SMC posterior samples through the SEIRD model to compute $\beta(t)$ for each draw.

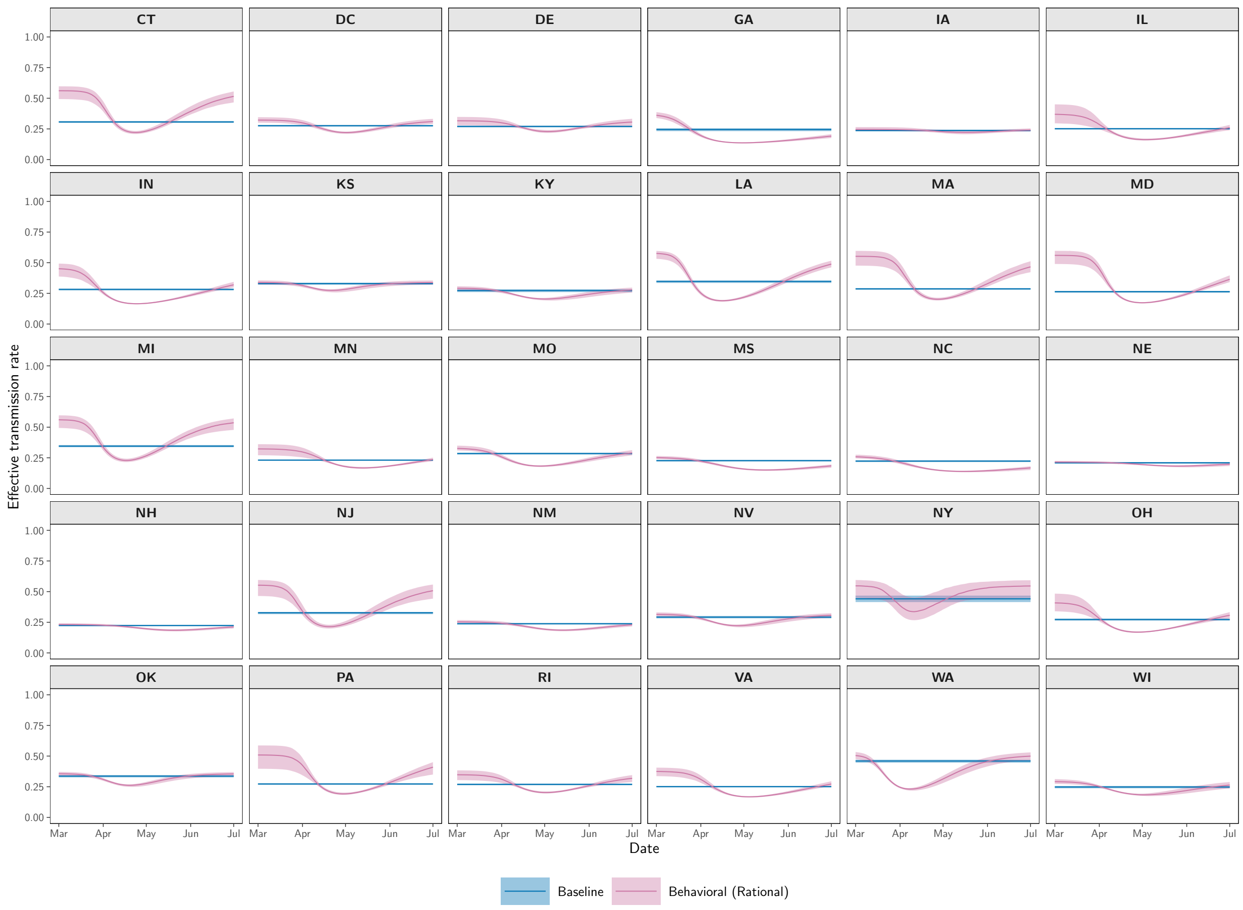

**Figure S11: Effective transmission rate $\beta(t)$ under the Behavioral (Rational) model.** Shown are posterior median trajectories together with 90% pointwise credible intervals, obtained by propagating weighted ABC–SMC posterior samples through the SEIRD model to compute $\beta(t)$ for each draw.

#### S7 Effective Reproduction Number

For each model and location, we compute the effective reproduction number $\mathcal{R}_e(t)$ as

$$\mathcal{R}_e(t) = \mathcal{R}_0(t)\,\frac{S(t)}{N},$$

where $S(t)$ is the susceptible population and $N$ is the total population, as defined in Eq. **2.7**.

To quantify uncertainty, we propagate posterior parameter uncertainty by drawing 1,000 weighted samples from each model's ABC–SMC posterior. For each sampled parameter set, we simulate the SEIRD model, extract the aligned trajectories $S(t)$ and $I(t)$, compute $\mathcal{R}_e(t)$, and summarize across the ensemble using the 5th, 50th, and 95th percentiles. These form the posterior median curves and 90% pointwise credible intervals shown in Figures S12, S13, and S14.

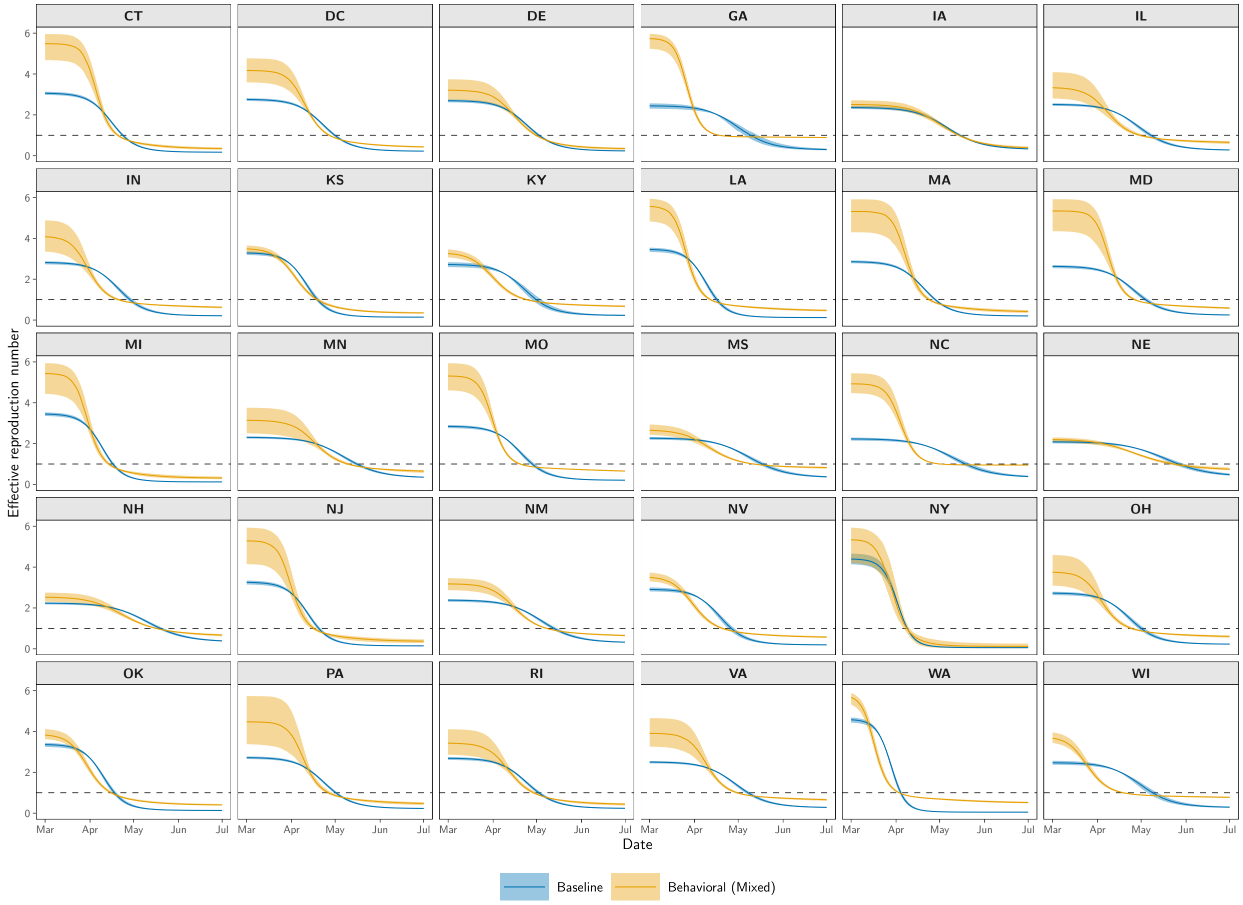

**Figure S12: Posterior effective reproduction number $\mathcal{R}_e(t)$ under the Behavioral (Mixed) model.** Posterior median curves and 90% pointwise credible intervals are shown for all locations.

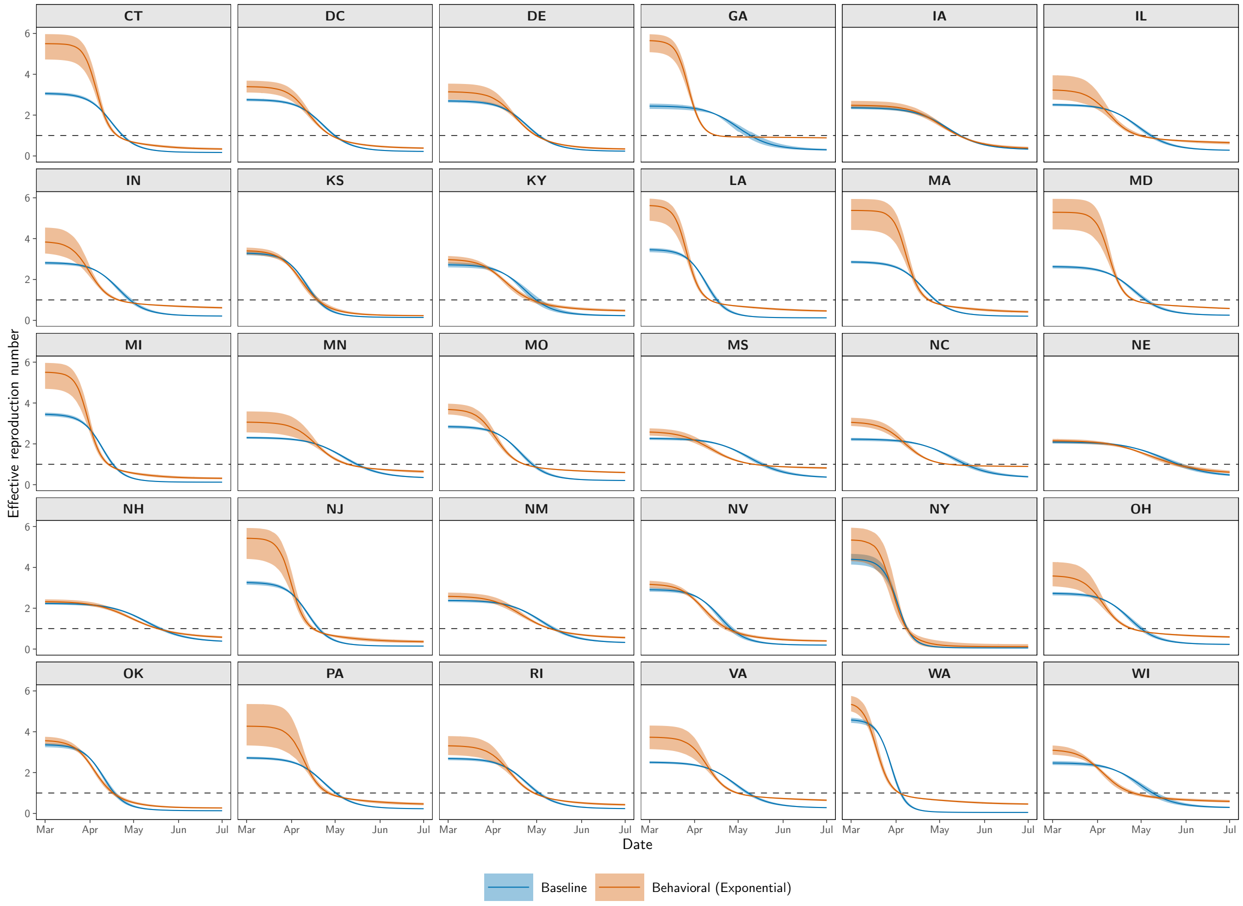

**Figure S13: Posterior effective reproduction number $\mathcal{R}_e(t)$ under the Behavioral (Exponential) model.** Posterior median curves and 90% pointwise credible intervals are shown for all locations.

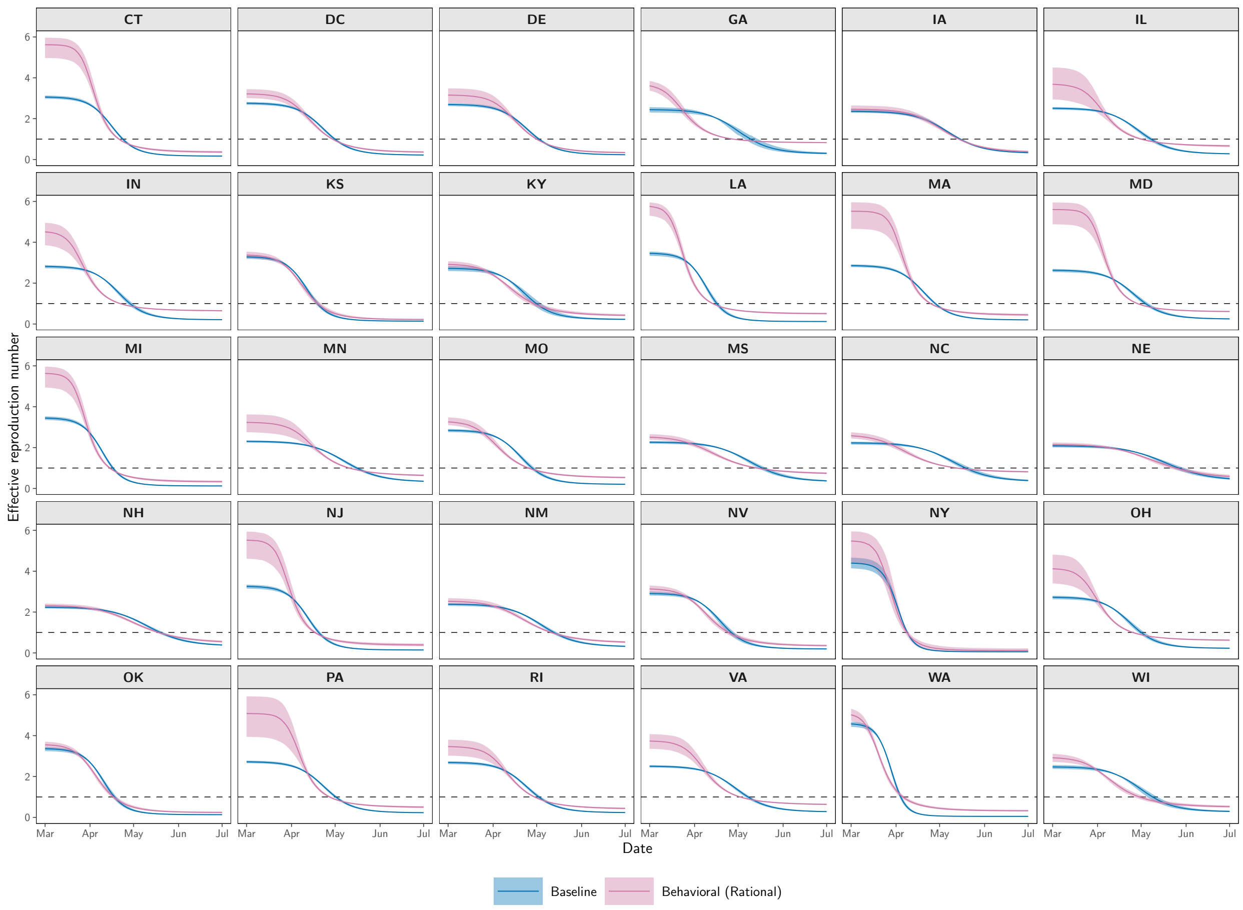

**Figure S14: Posterior effective reproduction number $\mathcal{R}_e(t)$ under the Behavioral (Rational) model.** Posterior median curves and 90% pointwise credible intervals are shown for all locations.
